# Contribution of leukocyte telomere length to major cardiovascular diseases onset: phenotypic and genetic insights from a large-scale genome-wide cross-trait analysis

**DOI:** 10.1101/2024.05.20.24307614

**Authors:** Jun Qiao, Qian Wang, Yuhui Zhao, Minjing Chang, Liuyang Cai, Feng Liu, Kaixin Yao, Leilei Zheng, Ning Tan, Pengcheng He, Anil G. Jegga, Siim Pauklin, Lei Jiang, Yining Yang, Yuliang Feng

## Abstract

Telomere shortening, a marker of cellular aging and genomic instability, has been epidemiologically linked to an increased risk of various cardiovascular diseases (CVDs). However, shared genetic determinants involved in these associations remain unclear. We composed an atlas of the shared genetic associations between leukocyte telomere length (LTL) and six major CVDs by investigating shared genetic elements, encompassing SNPs, genes, biological pathways, and protein targets with pleiotropic implications. Extensive genetic overlaps beyond genetic correlations were observed, but no causal relationships were established. We identified 248 independent pleiotropic genomic risk loci, implicating 50 unique genes in two or more trait pairs, especially the *SH2B3* gene, which was further validated by a proteome-wide Mendelian Randomization study. Functional analysis demonstrated a link to both DNA biosynthetic processes and telomere maintenance mechanisms. These findings suggest a genetic link between LTL and CVDs, highlighting a shared genetic basis crucial for developing future interventions and therapeutic targets.

## Introduction

Telomeres, DNA-protein complexes located at the ends of linear eukaryotic chromosomes, play a critical role in genome protection and act as indicators of biological aging.^1^. With each cell division, telomere progressively shortens due to the increased rate of somatic cell turnover and aging, eventually reaching a critical threshold known as the Hayflick limit. Beyond this point, DNA damage and cellular senescence begin, marking the onset of genomic instability. This process of telomere attrition is particularly relevant in cardiovascular diseases (CVDs), where the accumulation of senescent cells leads to tissue inflammation and matrix degradation. These changes contribute to the thinning of the fibrous cap, thereby heightening the risk of CVDs. Previous meta-analyses have identified an inverse association between leukocyte telomere length (LTL) and the risk of most CVDs, particularly coronary artery disease (CAD) and heart failure (HF), independent of conventional vascular risk factors, although the relationship with other CVDs remains more ambiguous^2–4^. This body of evidence highlighted the potential impact of LTL on the development and progression of various CVDs. Moreover, inflammation and oxidative stress central to the pathogenesis of CVDs further accelerate telomere shortening and cellular senescence. This complex interplay highlights the significant connection between LTL and CVDs, underscoring the need for further research into their joint mechanisms.

Previous epidemiological studies have demonstrated that telomere shortening was associated with an elevated risk of various CVDs. One plausible explanation is the overlap of genetic determinants for LTL and CVDs. LTL varies significantly among individuals from birth and throughout their lifespan, exhibiting high heritability with estimates ranging from 44% to 86%^5,6^. The largest genome-wide association study (GWAS) to date on LTL, using UK Biobank data, identified 138 associated loci, revealing links between genetically determined LTL^7^ and multiple CVD phenotypes, such as CAD. The recent accessibility of GWAS data for a diverse spectrum of CVD phenotypes offers a valuable opportunity to investigate the genetic basis between LTL and CVDs^8–13^. The shared genetic foundations may be understood as genetic variants influencing multiple complex phenotypic traits through both vertical and horizontal pleiotropy. Briefly, vertical pleiotropy emerges when a genetic variation affects one phenotype, which in turn influences the occurrence of another phenotype—a concept primarily addressed by Mendelian Randomization (MR) studies. Several previous MR studies have suggested that the associations of shorter LTL with CAD and Stroke were genetically causal, although evidence for an association with AF and HF was less certain, presenting confused results^14–17^. The causality of the relationships between LTL and other CVD phenotypes has not been adequately demonstrated. Conversely, horizontal pleiotropy occurs when a single genetic variation simultaneously impacts multiple phenotypes, which may highlight potential shared biological pathways among complex traits. Recent advances in genomics statistical tools have unveiled the vital role of horizontal pleiotropy beyond vertical pleiotropy in shared genetic foundations across complex traits, as highlighted in a study by Gong *et al*^18^. However, prior research on horizontal pleiotropy has primarily concentrated on exploring the relationship between LTL and severe mental disorders^19^. Therefore, in light of the widespread associations reported in epidemiological studies and the existing evidence of a shared genetic basis LTL and CVDs, the necessity for a comprehensive, large-scale analysis is clear.

Recognizing the knowledge gap, our study aims to provide a thorough analysis of shared genetic architectures between LTL and CVDs by encompassing the unprecedentedly large available GWAS datasets in individuals of European ancestry. First, by charting the landscape of genetic overlap beyond genetic correlation, we provide more granular insights into the unique and shared genetic architectures between LTL and CVDs. Then, we employed statistical techniques to capture diverse forms of genetic pleiotropy, followed by thorough analyses to link the genomic findings to biological pathways, yielding profound implications for conceptualizing shared genetic risk for both LTL and CVDs.

## Results

### Genetic overlap beyond genetic correlation between LTL and six major CVDs

Following the harmonization and filtering of SNPs shared across GWAS summary statistics, we employed cross-trait linkage disequilibrium (LD) score regression (LDSC) to calculate SNP-based heritability (*h^2^_SNP_*) and to assess genome-wide genetic correlation (*r_g_*) between LTL and six major CVDs. Univariate LDSC revealed that the estimated *h^2^_SNP_*for LTL was 2.76% (SE = 0.30%). On average, the estimated *h^2^_SNP_* was nearly threefold higher for AF (*h^2^_SNP_* =2.50%, SE = 0.33%), CAD (*h*^2^ = 3.25%, SE = 0.19%), and VTE (*h*^2^ =1.82%, SE = 0.23%), in comparison to HF (*h^2^_SNP_* = 0.80%, SE = 0.06%), PAD (*h^2^_SNP_* = 0.94%, SE = 0.13%), and Stroke (*h^2^_SNP_* = 0.60%, SE = 0.05%) (Supplementary Fig. 1a and Supplementary Table 2a). The results of bivariate LDSC indicated a range of weak to moderate genome-wide *r_g_* between LTL and CVDs, excluding LTL-AF. Notably, the most pronounced negative *r_g_*were observed for LTL-PAD (*r_g_* = −0.250, SE= 0.040, *P* = 3.83×10^−10^) and CAD (*r_g_* = −0.171, SE = 0.025, *P* = 4.65×10^−12^), while smaller but statistically significant *r_g_* were noted for Stroke (*r_g_* = −0.104, SE = 0.037, *P* = 4.60×10^−3^) and VTE (*r_g_* = −0.072, SE = 0.025, *P* = 4.20×10^−3^). LTL was only moderately genetically correlated to HF (*r_g_* = −0.145, SE = 0.037, *P* = 8.20×10^−5^). In contrast, no significant *r_g_* was observed between LTL and AF (Supplementary Fig. 1b and Supplementary Table 2b). Although genome-wide *r_g_* offered valuable insight into the genetic overlap between phenotypes, it could not distinguish genetic overlap resulting from a mixture of concordant and discordant effects from the absence of genetic overlap, potentially yielding an estimated *r_g_* near zero in both scenarios. Therefore, using multiple methods with different model assumptions to identify and understand this “missing dimension” of genetic overlap was essential for comprehensively characterizing the shared genetic foundations across phenotypes.

**Figure 1:**
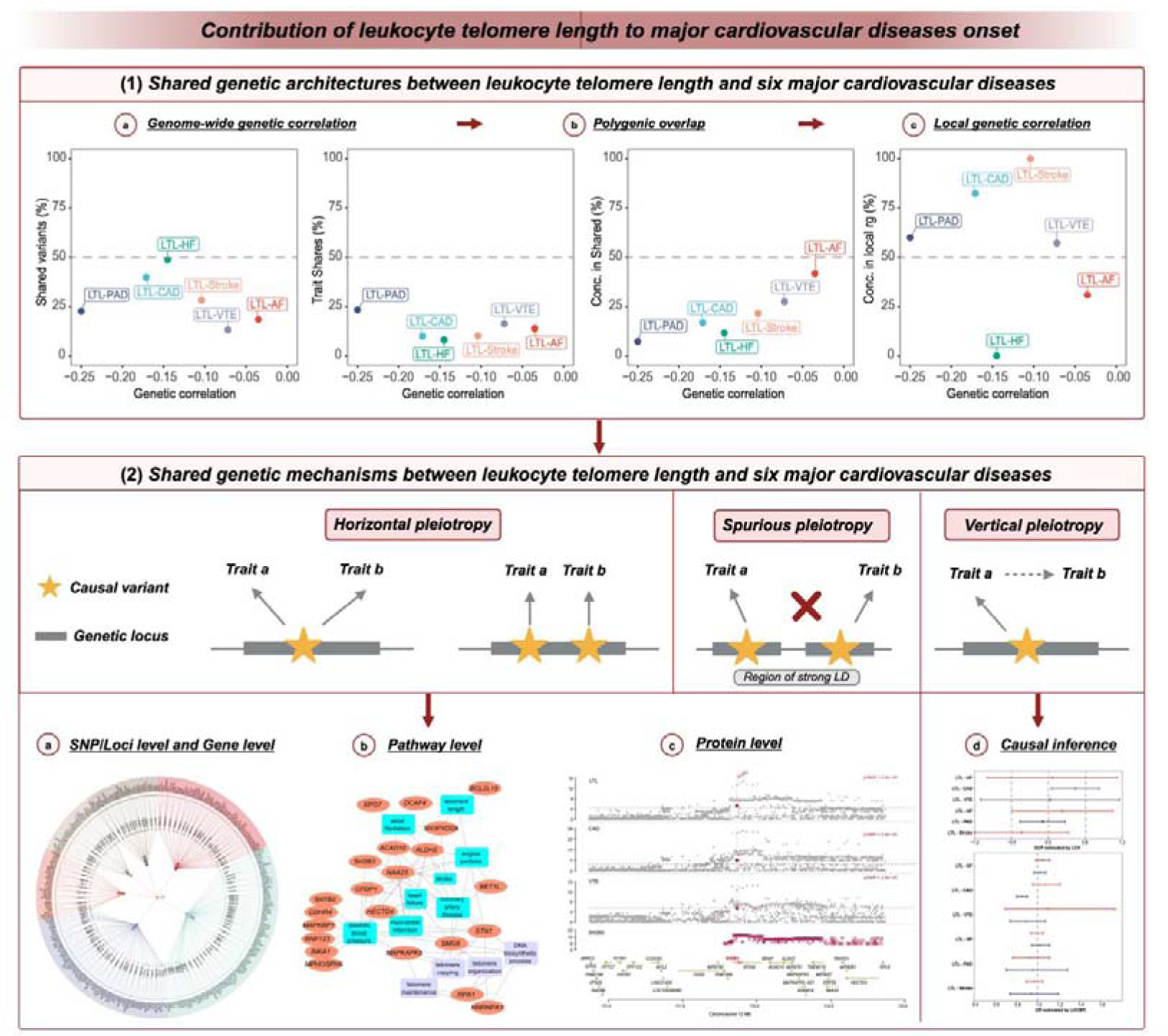
Schematic representation of analyses performed for leukocyte telomere length and six major cardiovascular diseases in the current study. This figure demonstrates the comprehensive pleiotropic analysis conducted for LTL and six major CVDs from multiple perspectives within this study. We first investigated the shared genetic architectures between LTL and six major CVDs by assessing pairwise genetic overlap beyond correlation. Extensive analyses were then conducted to investigate two types of pleiotropy: horizontal pleiotropy, whereby causal variants for two traits colocalize in the same locus, and vertical pleiotropy, whereby a variant exerts an effect on one trait through another. Notably, spurious pleiotropy was excluded from the analysis, whereby causal variants for two traits fall into distinct loci but are in LD with a variant associated with both traits. Therefore, We applied Mendelian randomization to evaluate the pairwise causal associations between LTL and the CVDs, primarily elucidating the contributions from vertical pleiotropy. We used novel statistical tools to capture horizontal pleiotropy by characterizing the shared loci and their implications on genes, tissues, biological functions, and protein targets. This comprehensive pleiotropic analysis allowed us to construct an atlas of the shared genetic associations, enhancing our understanding of the complex interactions between LTL and cardiovascular health. The diagram was generated using BioRender (www.biorender.com) and has been included with permission for publication. LTL, leukocyte telomere length; AF, Atrial fibrillation; CAD, Coronary artery disease; VTE, Venous thromboembolism; HF, Heart failure; PAD, Peripheral artery disease.

Despite minimal *r_g_* estimated by LDSC, the causal mixture modeling approach (MiXeR) was then applied to elucidate extensive genetic overlap beyond genetic correlation by determining the number of overlapping variants between LTL and six major CVDs, irrespective of the direction of their effects. Univariate MiXeR revealed that LTL exhibited a lower degree of polygenicity (N = 0.380K, SD = 0.026K). Among the six major CVDs, HF (N = 2.305K ‘causal’ variants explaining 90% of HF’s *h^2^_SNP_*, SD = 0.213K) was the most polygenic, followed by CAD (N = 1.528K, SD = 0.311K) and Stroke (N = 1.055K, SD = 0.117K). VTE, PAD, and AF demonstrated lower polygenicity, associated with 0.308K to 0.504K variants at 90% *h*^2^ (Supplementary Table 3a). These findings highlighted a pattern of polygenicity distinct from *h^2^_SNP_*estimates.

The result of bivariate MiXeR revealed substantial but distinct patterns of polygenic overlap between LTL and CVDs. Given the low polygenicity of LTL and these CVD phenotypes (including AF, PAD, and VTE), LTL was found to share less proportion of causal variants with these CVDs, ranging from 18.30% in AF to 22.63% in PAD. However, relatively large genetic overlaps were also observed between LTL and these CVDs (Fig. 2a, Supplementary Fig. 2, Supplementary Table 3b). For example, polygenic overlap between LTL and PAD was particularly striking (Dice coefficientL=L0.229, SD = 0.015), with 0.086K (SDL=L0.007K) shared variants, representing 22.63% LTL-influencing variants and 23.24% PAD-influencing variants, consistent with the strongest negative genome-wide genetic correlation (*r_g_* = −0.223, SE = 0.014) and genetic correlation of shared variants (*r_g_s* = −0.970, SE = 0.022). A total of 0.069K (SDL=L0.019) variants were estimated to be shared between LTL and AF, representing 18.30% LTL-influencing variants and 13.79% AF-influencing variants, despite weak negative genetic correlation. This pattern of extensive genetic overlap but weak *r_g_*indicated a predominance of mixed effect directions, supported by the MiXeR-estimated proportion of shared ‘causal’ variants with concordant effects (0.416, SD = 0.029). Considering the low polygenicity of LTL and high polygenic diseases such as CAD, HF, and Stroke, significant disparities were observed in the number of shared and unique “causal” variants. In particular, MiXeR estimated that of the 0.380K LTL-influencing variants, 48.69%, 39.82%, and 28.07% also influence HF, CAD, and Stroke, respectively. For example, LTL and HF shared the largest number of variants (N = 0.185K, SD = 0.024K), with many more unique variants of HF (N = 2.12K, SD = 0.208K) than unique variants of LTL (0.195K, SD = 0.030K), representing 48.69% LTL-influencing variants and 8.02% HF-influencing variants. While they were moderately correlated at the genome-wide level (*r_g_* = −0.180, SE = 0.017), shared variants were strongly correlated (*r_g_s* = −0.913, SE = 0.087). A similar, although less pronounced, relationship was evident in LTL-CAD and LTL-Stroke. Furthermore, the overlapping variants demonstrated a low level of effect direction concordance, highlighting the prevalence of mixed effect directions between LTL and CVDs. These observations suggested that the extent of polygenic overlap between LTL and CVDs were likely underestimated by genome-wide genetic correlations (Fig. 2b).

**Figure 2:**
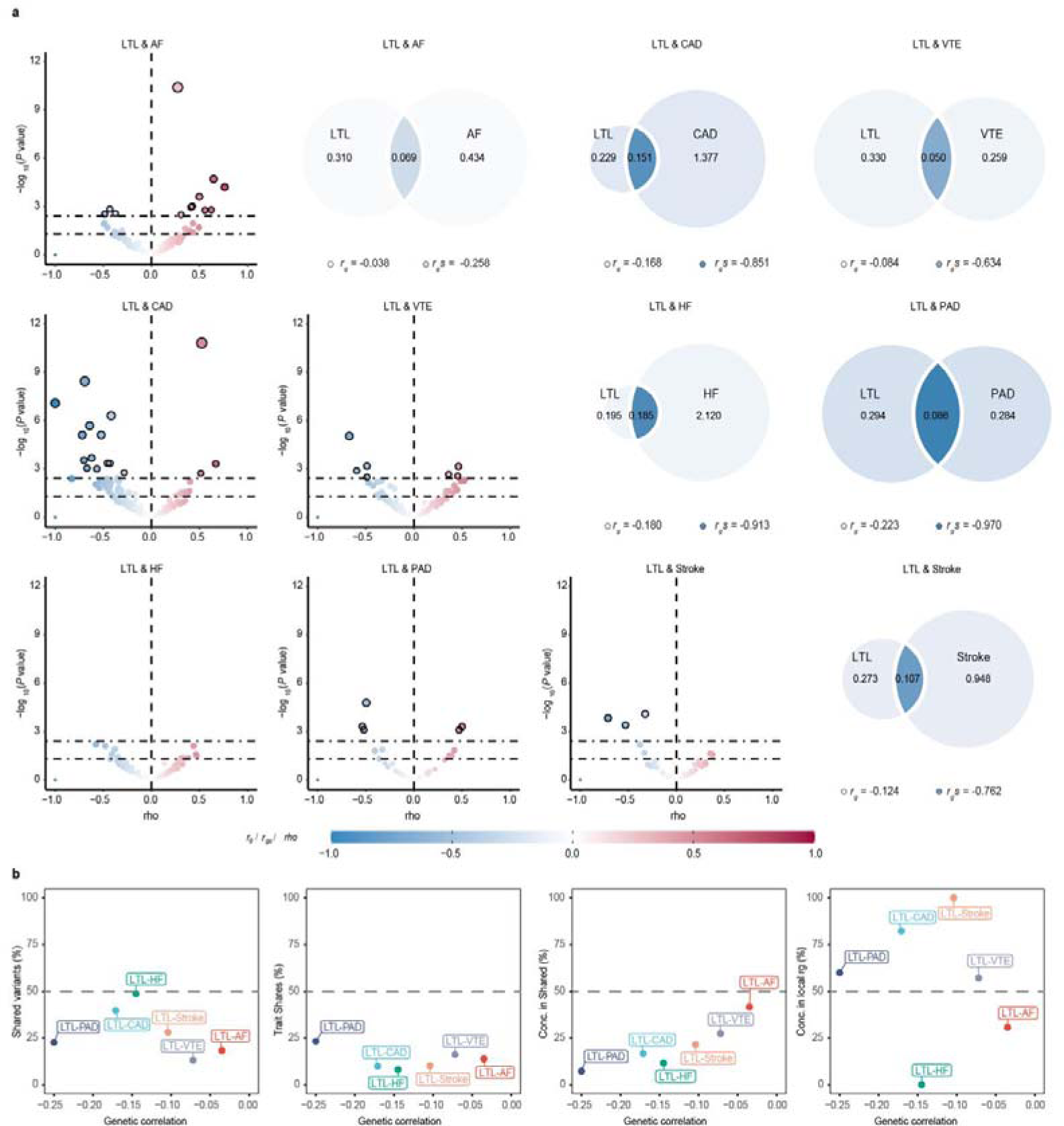
Genetic overlap between leukocyte telomere length and six major cardiovascular diseases beyond genome-wide genetic correlation. (a) MiXeR-modeled genome-wide genetic overlap and genetic correlations (top right) and LAVA local correlations (bottom left) between LTL and six major CVDs. Top right: MiXeR Venn diagrams showing the number (in thousands) of estimated ‘causal’ variants that are unique to LTL (left circle), six major CVDs (non-overlapping part of the right circle) or shared between LTL and six major CVDs (overlapping part of circles). Genome-wide genetic correlation (*r_g_*) and genetic correlation of shared variants (*r_g_s*) are represented by the color of the trait-specific (*r_g_*) and shared regions (*r_g_s*), respectively. The circle size represents the extent of polygenicity of each trait, with larger circles corresponding to greater polygenicity and vice versa. Bottom left: Volcano plots of LAVA local genetic correlation coefficients (rho, y-axis) against −log10 (p-values) for each pairwise analysis per locus. Larger dots with black circles represent significantly correlated loci after FDR correction (FDR < 0.05). MiXeR estimated *r_g_* and *r_g_s*, and LAVA estimated rho are represented on the same blue to red color scale. Note that the volcano plots were plotted at p-values truncated by 1L×L10^−F12^ for better visualization, thus excluding a region (LD block 1,581 on chromosome 10, ranges from 104,206,838 to 106,142,283) influencing LTL - CAD from volcano plots of LAVA. (b) Genetic correlation estimated by LDSC (x-axis) against the percentage of LTL variants that are shared with CVDs as estimated by MiXeR (first plot), the percentage of CVD variants that are shared with LTL (second plot), and the percentage of CVD variants that are shared with LTL that have concordant effect directions (third plot). The fourth plot shows the percentage of local genetic correlations from LAVA with concordant effect directions on the y-axis. LTL, leukocyte telomere length; AF, Atrial fibrillation; CAD, Coronary artery disease; VTE, Venous thromboembolism; HF, Heart failure; PAD, Peripheral artery disease.

Local genetic correlations provide a more effective means of capturing genetic associations with mixed effect directions. Specifically, a pair of traits may display no genome-wide *r_g_* due to an equal number of positive and negative (opposite effect directions) local genetic correlations with comparable magnitudes. To prevent the potential masking of local genetic correlations when evaluating *r_g_* at the genome-wide level, we applied Local Analysis of [co]Variant Annotation (LAVA) to perform local genetic correlations (loc-*r_g_s*) between LTL and six major CVDs at loci, where both phenotypes had heritability estimates significantly different from zero (Supplementary Table 4). Overall, 45 local genomic regions were found significant for bivariate analysis after correcting for multiple testing using FDR (FDR < 0.05, Fig. 2a and Supplementary Table 5), with 62% and 38% of the partitions showing negative and positive loc-*r_g_s*, respectively. Corroborating the MiXeR findings, LAVA estimated correlated loci of LTL-PAD (2 positively correlated and 3 negatively correlated loci), LTL-VTE (3 positively correlated and 4 negatively correlated loci), and LTL-AF (9 positively correlated and 4 negatively correlated loci), adding further support for a shared genetic basis. Local correlations for LTL-CAD, comprising 14 negatively and 3 positively correlated loci, and LTL-Stroke, with 3 negatively correlated loci and none positively correlated, were inconsistent with the mixed effect directions estimated by MiXeR. The absence of significant loci between LTL and HF may be attributed to LAVA’s propensity for identifying loci with extreme correlations, in contrast to MiXeR, thereby highlighting loci more likely to be statistically significant.

Interestingly, our investigation also identified a solitary region (LD block 1,841 on chromosome 12, ranges from 111,592,382 to 113,947,983) displaying significant correlations for a majority of the trait pairs, with uniform negative correlation values between −0.530 and −0.692. Subsequent analyses utilizing Hypothesis Prioritisation in Multi-trait Colocalization (HyPrColoc) revealed robust colocalization evidence for this locus between LTL and all CVDs, excluding AF, HF and PAD, with a posterior probability (PP) higher than 0.7, which encompassed the shared causal SNP (rs10774625, an intronic variant of the ataxin 2 (*ATXN2*) gene on 12q24.12). The *ATXN2* rs10774625 polymorphism has been associated with various CVDs, notably CAD, alongside cardiometabolic markers such as blood pressure and blood lipids^20,21^.

### The Causal inference between LTL and six major CVDs

Despite these findings substantiating a shared genetic foundation between LTL and six major CVDs, there was uncertainty in relation to whether the complex interplay predominantly reflected horizontal pleiotropy or potentially involved a causal relationship (referred to as ‘vertical pleiotropy’). Mendelian randomization (MR) harnesses vertical pleiotropy to deduce potential causal relationships, excluding any SNPs indicative of horizontal pleiotropy. Therefore, latent causal variable (LCV) analysis was utilized to elucidate the possible causal relationships underlying the genetic correlations observed. Notably, none of the trait pairs demonstrated tendencies indicative of partial genetic causation (Supplementary Fig. 3a, Supplementary Table 6a). To ascertain the reproducibility of the partial causal associations between trait pairs, we utilized the Latent Heritable Confounder Mendelian Randomization (LHC-MR) approach, taking into account factors such as sample overlap, bidirectional causal associations, and unobserved heritable confounders. Consistently, no evidence was found to support a putative causal effect of LTL on CVDs (Supplementary Fig. 3b, Supplementary Table 6b). Only the possible weak positive causal impact of CAD on LTL was observed, yet lacked confirmation from conventional bidirectional MR analyses (Supplementary Table 6c), emphasizing the need for cautious interpretation. Overall, MR analysis revealed that the shared genetic foundation between LTL and CVDs cannot be ascribed to vertical pleiotropy.

### Pleiotropic genomic loci identified for LTL and CVDs

The observed comorbidity between LTL and six major CVDs suggests that instead of a predominance of trait-specific risk variants, there may be a set of pleiotropic variants influencing the risk of both LTL and CVDs (i.e., horizontal pleiotropy). To investigate this, we employed the Pleiotropic Analysis under a Composite Null Hypothesis (PLACO) to pinpoint potential pleiotropic variants shared between LTL and CVDs, resulting in the identification of 12,604 SNPs comprising 10,008 unique variants. Functional Mapping and Annotation (FUMA) further delineated 248 independent genomic risk loci as pleiotropic, spanning 122 unique chromosomal regions (Fig. 3, Supplementary Fig 4, Supplementary Table 7). Among these, 194 loci were associated with LTL and 80 loci with CVDs. In aggregate, 32 loci overlapped between LTL and CVDs, accounting for 16.49% and 40.00% of the total number of loci linked to these respective categories. A total of 188 pleiotropic loci exhibited genetic signals for multiple trait pairs, with 74 of them (39.36%) spanning 16 unique chromosomal regions, demonstrating this phenomenon in over half of the investigated trait pairs. For example, the pleiotropic locus 16q22.1 (mapped gene: *TMED6*) was jointly associated with LTL and all CVDs. A mixture of concordant and discordant allelic effects existed in these pleiotropic loci. Remarkably, the selected effect alleles at the top SNPs within or near 117 loci (47.2%) exhibited inconsistent effects on two traits within a pair of traits. Essentially, these variants may concurrently increase LTL and diminish the risk of developing CVDs, aligning with their robust genome-wide genetic correlation. Conversely, the remaining SNPs demonstrated concordant associations with both LTL and CVDs, implying that these SNPs might influence the risk of both traits in the same direction.

**Figure 3:**
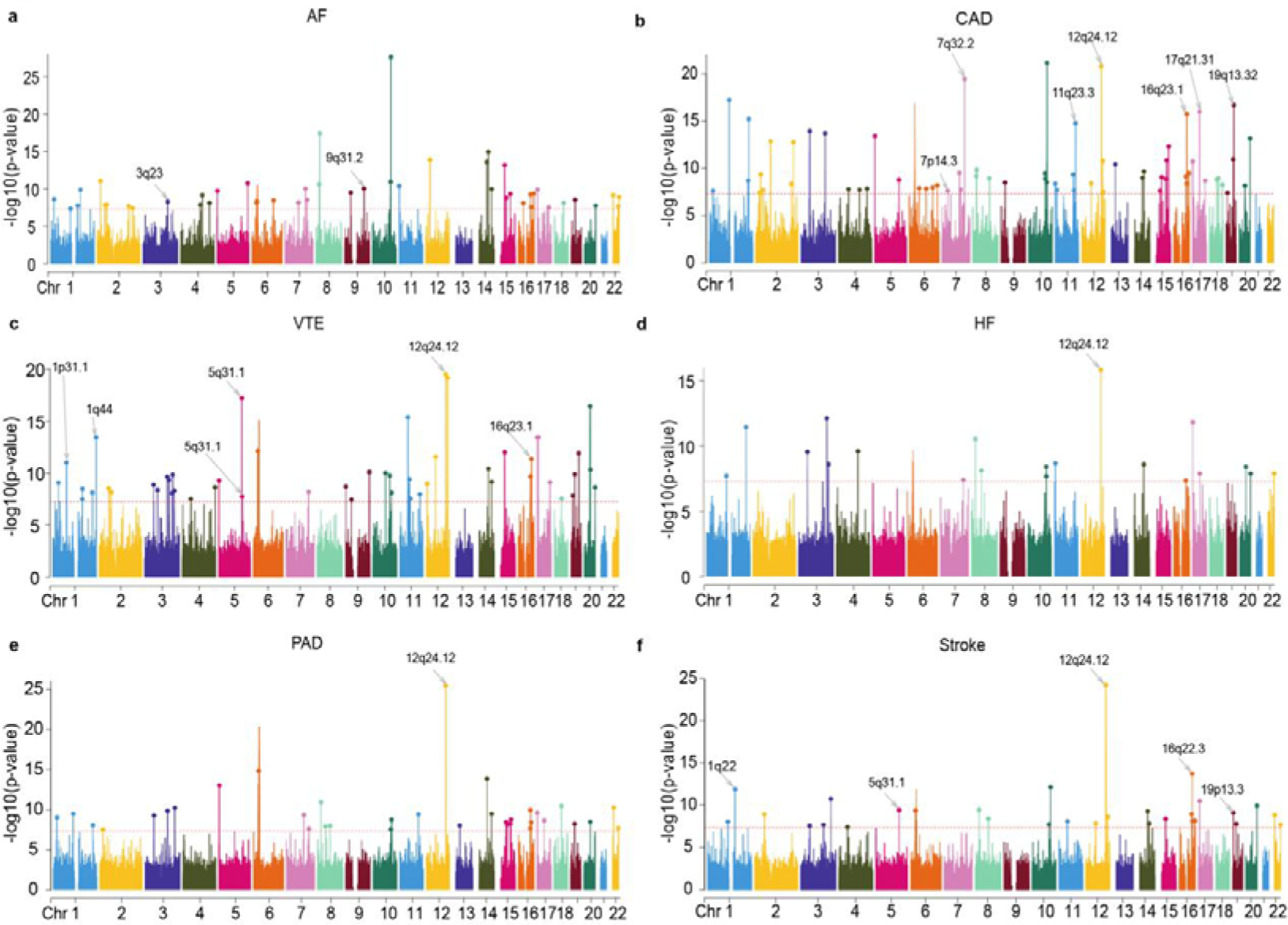
Manhattan plots for the PLACO results of leukocyte telomere length and six major cardiovascular diseases. The x-axis reflects the chromosomal position, and the y-axis reflects negative log10 transformed *P*-values for each SNP. The horizontal dashed red line indicates the genome-wide significant *P*-value of −log10 (5×10^−8^). The independent genome-wide significant associations with the smallest P-value (Top lead SNP) are encircled in a colorful circle. Only SNPs shared across all summary statistics were included. Labels are the chromosome regions where genomic risk loci with strong evidence for colocalization (PP.H4 > 0.7) are located. LTL, leukocyte telomere length; AF, Atrial fibrillation; CAD, Coronary artery disease; VTE, Venous thromboembolism; HF, Heart failure; PAD, Peripheral artery disease.

ANNOVAR category annotation of candidate SNPs shared between LTL and CVDs revealed that 67 (27.0%) were in intergenic regions, 114 (45.9%) were in intronic regions, and only 14 (5.6%) were in exonic regions. For example, the index SNP rs1566452 at 16q22.1 locus (*P_PLACO_* =L4.46×10^−8^ for LTL-HF) was associated with artery coronary and artery tibial eQTLs (*P_Artery_Coronary_*= 4.06×10^−5^, *P_Artery_Tibial_*= 3.32×10^−10^, Supplementary Table 9) for WW domain-containing E3 ubiquitin protein ligase 2 (*WWP2*) gene encoding one of the E3 ubiquitin ligases, which critically participate in the development and progression of cardiovascular diseases^22^. Besides, numerous ubiquitin E3 ligases have also been documented to promote the degradation of human telomerase reverse transcriptase (hTERT), thereby reducing telomerase activity and potentially leading to decreased telomere length^23^. Furthermore, we identified 21 top SNPs with combined annotation-dependent deletion (CADD) scores exceeding 12.37, and 7 mRNA exonic variants had higher CADD scores, which was indicative of potentially deleterious effects. Notably, rs11556924 within the zinc finger C3HC-type containing 1 (*ZC3HC1*) gene represents an exonic non-synonymous variant with a CADD score of 28 (variants with scores surpassing 20 are predicted to be among the 1.0% most deleterious substitutions in the human genome). Furthermore, nine SNPs were assigned RegulomeDB scores of 1f, 1d, or 1a, indicating a likely influence on binding sites. We followed up this finding using the GTEx database to investigate the gene regulatory effects. For example, rs11779558 at 8p21.3 locus was significantly associated with eQTL functionality in artery tibial (*P_Artery_Tibial_* = 5.79×10^−5^) for exportin 7 (*XPO7*) gene.

Colocalization analysis further revealed 22 out of 248 potential pleiotropic loci with PP.H4 greater than 0.7, wherein 14 top SNPs at the corresponding loci were identified as candidate-shared causal variants (Fig. 3, Supplementary Fig. 5, Supplementary Table 7). Notably, the 12q24.12 locus, identified as pleiotropic for all correlated trait pairs except for LTL-AF, exhibited strong evidence of colocalization between these trait pairs (PP.H4 ranging from 0.729 to 0.998). HyPrColoc analysis further revealed strong colocalization evidence for this locus between LTL and all CVDs except AF and PAD, with a PP exceeding 0.7, which encompassed rs10774625 (an intronic variant of the *ATXN2* gene on 12q24.12) as the potential shared causal variant. Moreover, 40 pleiotropic loci were identified with PP.H3 exceeding 0.7, indicating the possibility of different causal variants within these loci.

### Pleiotropic genes associated with LTL and multiple CVDs

Despite the success of the above analyses in identifying disease risk loci, the biological significance of most identified variants remains unknown. To achieve a more comprehensive understanding of how genetic variation influences disease risk, we adopted approaches integrating SNPs across a spectrum of association significance to construct a cohort of predicted genes that could subsequently be mapped to functional pathways for analysis. We employed two distinct strategies for mapping SNPs to genes: Firstly, a genome-wide gene-based association study (GWGAS) in MAGMA and positional mapping in FUMA were utilized, mapping SNPs to genes based on their physical position in the genome. Secondly, an eQTL-informed GWGAS in e-MAGMA and eQTL mapping in FUMA were employed to map SNPs to genes through their eQTL associations.

MAGMA analysis, utilizing 557 potential pleiotropic genes located within or overlapping with 248 pleiotropic loci, identified 478 significant pleiotropic genes (323 unique), in which 244 genes were detected in two or more trait pairs (Fig. 4, Supplementary Table 11). For example, *SH3PXD2A*, *SH2B3*, *BRAP*, *ATXN2*, *PTPN11*, *NAA25*, *ALDH2*, and *ACAD10* were identified as significant pleiotropic genes in five pairs of traits. Remarkably, seven of eight genes (excluding *SH3PXD2A*) were located on the 12q24.12 locus, identified in all trait pairs except for LTL-AF. Of the pleiotropic genes identified, 98 (20.50%) were novel for LTL and 258 (53.97%) for CVDs. Only one pleiotropic gene, CD19 molecule (*CD19*), had not previously been reported to be associated with both traits. Furthermore, 472 genes (98.74%) identified by MAGMA were confirmed by FUMA positional mapping (Supplementary Table 9).

**Figure 4:**
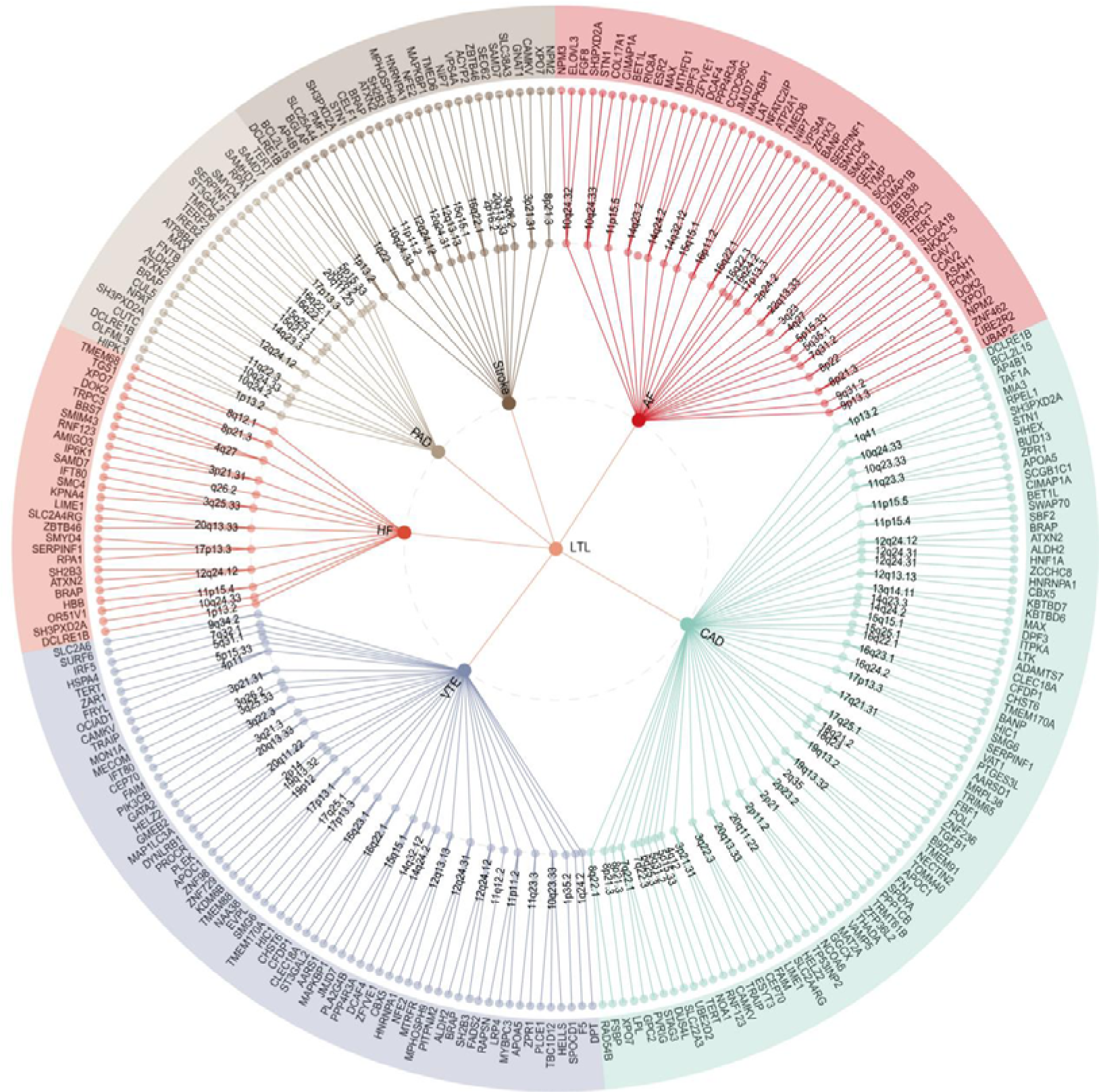
The Overall landscape of the pleiotropic associations across leukocyte telomere length and six msjor cardiovascular diseases. A circular dendrogram showing the shared genes between LTL (center circle) and each of six CVDs (first circle), resulting in six pairs. A total of 248 shared loci were identified across six trait pairs, mapped to 478 significant pleiotropic genes (323 unique) identified by multimarker analysis of GenoMic annotation (MAGMA). For the trait pairs with more than three pleiotropic genes, we only showed the top 3 pleiotropic genes according to the prioritization of candidate pleiotropic genes (fourth circle). LTL, leukocyte telomere length; AF, Atrial fibrillation; CAD, Coronary artery disease; VTE, Venous thromboembolism; HF, Heart failure; PAD, Peripheral artery disease.

To pinpoint tissues potentially integral to the biological processes of LTL and six major CVDs, we utilized LDSC-SEG for tissue-specific enrichment analysis using single trait GWAS summary statistics. Regarding multi-tissue gene expression, we found that expressions of LTL-associated loci were significantly enriched in spleen tissues, surpassing an FDR > 0.05 threshold (Fig. 5, Supplementary Table 14). Additionally, AF showed significant enrichment in heart-related tissues, including heart left ventricle and heart atrial appendage, while CAD demonstrated enrichment in artery-related tissues, such as artery tibial, artery aorta, and artery coronary. Conversely, no significant tissue-specific enrichment was observed for VTE, HF, PAD, and Stroke. These findings were corroborated by multi-tissue chromatin interaction results.

**Figure 5:**
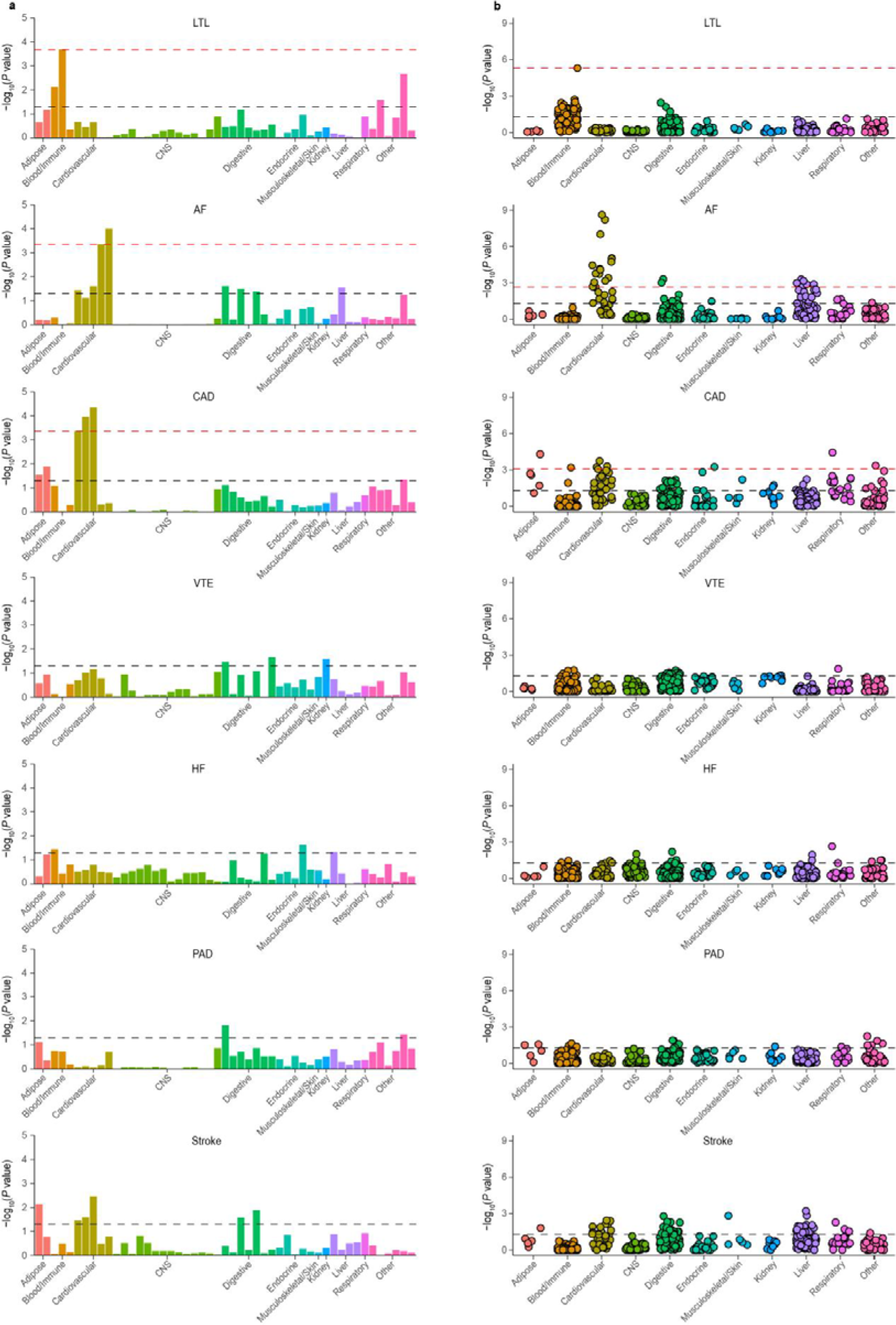
The results of multiple-tissue analysis using gene expression data and chromatin data for leukocyte telomere length and six cardiovascular diseases. **(a)** Tissue type-specific enrichment of single nucleotide polymorphism (SNP) heritability for LTL and CVDs in 49 tissues from GTEx v8 estimated using stratified LDSC applied to specifically expressed genes (LDSC-SEG). Each bar represents a tissue from the GTEx dataset. The x-axis reflects tissue types, and the y-axis reflects negative log10 transformed P-values. **(b)** Each point represents a peak for DNase I hypersensitivity site (DHS) or histone marks (including H3K27ac, H3K36me3, H3K4me1, H3K4me3, and H3K9ac) in a tissue or cell type. Tissues or cell types were classified into 11 distinct categories, i.e., ‘Adipose,’ ‘Blood/Immune,’ ‘Cardiovascular,’’ ‘CNS,’ ‘Digestive,’ ‘Endocrine,’ ‘Musculoskeletal/Skin,’ ‘kidney,’ ‘Liver,’ ‘Respiratory,’ and ‘Other.’ The black dotted line represents the significance threshold of *P* < 0.05, and the red line indicates the significant P-value after conducting FDR correction (FDR < 0.05). The color of the dots in a and b indicates different tissue types. LTL, leukocyte telomere length; AF, Atrial fibrillation; CAD, Coronary artery disease; VTE, Venous thromboembolism; HF, Heart failure; PAD, Peripheral artery disease.

Given that MAGMA assigns SNPs to the nearest genes based on arbitrary genomic windows, and considering that the effects of a locus don’t always operate through the nearest gene, a critical need remains to functionally link SNPs to genes (e.g., through genetic regulation) to enhance our understanding of potential underlying mechanisms. Consequently, e-MAGMA was conducted to uncover functional gene associations potentially overlooked by the proximity-based SNP assignment in MAGMA, thereby illuminating alternative causal pathways from SNPs to traits. E-MAGMA analysis revealed 1,844 significant tissue-specific pleiotropic genes (419 unique) after Bonferroni correction, each strongly enriched in at least one tissue (Supplementary Table 15). Of these, 918 tissue-specific pleiotropic genes (75 unique) were significantly identified across multiple trait-related tissues in at least two trait pairs. For example, *MAPKAPK5*, *TMEM116*, *HECTD4*, *ALDH2*, and *ACAD10*, all located on the 12q24.12 locus, were recognized as significant tissue-specific pleiotropic genes in all trait pairs except for LTL-AF. Notably, transmembrane protein 116 (*TMEM116*) gene was found to be highly tissue-specific, showing enrichment in eight trait-related tissues, including artery tibial, artery coronary, adipose visceral (omentum), adipose subcutaneous, heart atrial appendage, heart left ventricle, liver, and whole blood. *TMEM116* encoded a transmembrane protein involved in blood coagulation and had been identified as a potential risk gene for coronary atherosclerosis in previous studies^24^. However, its relationship with LTL remained less understood. In comparison with the transcriptome-wide association study (TWAS) results for single-trait GWAS, we identified 415 tissue-specific pleiotropic genes as novel for LTL and 851 for CVDs (Supplementary Table 16). Finally, we successfully replicated 1,117 genes (60.57%) using FUMA eQTL mapping (Supplementary Table 9).

Finally, 289 pleiotropic genes (207 unique) were jointly identified by MAGMA and e-MAGMA analysis, in which 50 unique genes were detected in 2 or more trait pairs, further suggesting the tissue specificity of these pleiotropic genes (Supplementary Table 11). For example, *ACAD10* (12q24.12), *ALDH2* (12q24.12), *HECTD4* (12q24.12), *MAPKAPK5* (12q24.12), *NAA25* (12q24.12), *SH2B3* (12q24.12), *TMEM116* (12q24.12), *SERPINF1* (17p13.3), *TMED6* (16q22.1), and *XPO7* (8p21.3) were identified as significant pleiotropic genes in more than half of the trait pairs. Remarkably, two of ten genes (including *ALDH2* and *ACAD10*) were located at the 12q24.12 locus, identified in five pairs of traits except for LTL-AF. Acyl-CoA dehydrogenase family member 10 (*ACAD10*), a gene encoding an enzyme crucial for fatty acid beta-oxidation in mitochondria, critically regulates cellular lipid synthesis with significant expression in the human brain^25^. Previous data show that homozygous loss of function of *ACAD10* results in perturbed lipid synthesis that potentially influences the development of CVDs, whereas common gene variants have been associated with CAD, Stroke, and hypertension (a common CVD risk factor). Additionally, aldehyde dehydrogenase 2 family member (*ALDH2*) gene encoding a vital mitochondrial enzyme critical for cardiac function, has been associated with exacerbated myocardial remodeling and contractile dysfunction in aging. This association is possibly through mitochondrial damage mediated by the AMPK/Sirt1 pathway.

### Shared biological pathways between LTL and six major CVDs

To investigate the concept that a group of genes might work collectively to fulfill specific biological functions through shared pathways or functional category enrichments, we employed various analytical strategies, including gene-set analysis of genes identified via MAGMA and functional enrichment analysis targeting tissue-specific genes. After rigorously adjusting for 7,744 gene sets (biological processes sets from the Molecular Signatures Database (MSigDB, v.2023.1; C5: GO BP)), we noted minimal overlap in gene sets between LTL and six major CVDs. Only three gene sets associated with chromatin organization, the negative regulation of nucleobase-containing compound metabolic processes, and the negative regulation of miRNA maturation were enriched across several trait pairs, and no gene set demonstrated significant enrichment across more than one trait (Supplementary Table 17a). Remarkably, massive genes shared between LTL and AF were most significantly linked to the ‘negative regulation of the nucleobase-containing compound metabolic process.’ Subsequently, we identified several biological processes overrepresented among the 50 pleiotropic genes detected by MAGMA and e-MAGMA analysis in 2 or more trait pairs shared between LTL and CVDs using the ToppGene Functional Annotation tool (ToppFun) (Supplementary Fig. 6, Supplementary Table 17b). All identified significant biological processes, with the exception of the DNA biosynthetic process, were directly associated with telomere maintenance processes such as ‘telomere maintenance via telomerase,’ ‘telomere maintenance via telomere lengthening,’ ‘telomere capping,’ and ‘telomere organization.’ Interestingly, the nucleobase-containing compound metabolic process identified in the MAGMA gene-set analysis encompassed both DNA biosynthetic processes and telomere maintenance mechanisms, implying a key role in shared biological pathways between LTL and CVDs.

### Shared causal proteins between LTL and six major CVDs

We undertook a comprehensive analysis of the Mendelian randomization (MR) associations between 1,922 unique proteins and the risk of LTL and six major CVDs using the summary data-based Mendelian randomization (SMR), each protein with index cis-acting variants (cis-pQTL) obtained from the UK Biobank Pharma Proteomics Project (UKB-PPP). Following the exclusion of associations failing the HEIDI test, and after conducting sensitivity analysis with multi-SNPs-SMR and applying multiple testing corrections via Bonferroni adjustment, the genetically predicted levels of 85 proteins were found to be significantly associated with the risk of LTL and CVDs (Fig. 6, Supplementary Table 18). Specifically, 12, 9, 26, 24, 2, 6, and 6 proteins were significantly associated with LTL, AF, CAD, VTE, HF, PAD, and Stroke, respectively. Notably, SH2B3 emerged as significantly associated with both LTL-CAD and LTL-VTE, also showing strong colocalization evidence in HyPrColoc analysis, with rs10774625 pinpointed as a shared causal variant. The index SNP rs10774625, located at the 12q24.12 locus (an intronic variant of the *ATXN2* gene), was associated with eQTLs in whole blood (*P_Whole_Blood_* = 3.48×10^−4^) and was also linked to pQTLs in whole blood (*P_Whole_Blood_*= 1.08×10^−3^) for the SH2B adaptor protein 3 (SH2B3). SH2B3 acted as an adaptor protein, playing a crucial role in negatively regulating cytokine signaling and cell proliferation. Prior research indicated that SH2B3 was associated with longevity, with missense alleles within SH2B3 influencing life expectancy through predisposition to cardiovascular events^26^. Enhancing SH2B3 function has demonstrated therapeutic potential in hypertension and other cardiovascular conditions.

**Figure 6:**
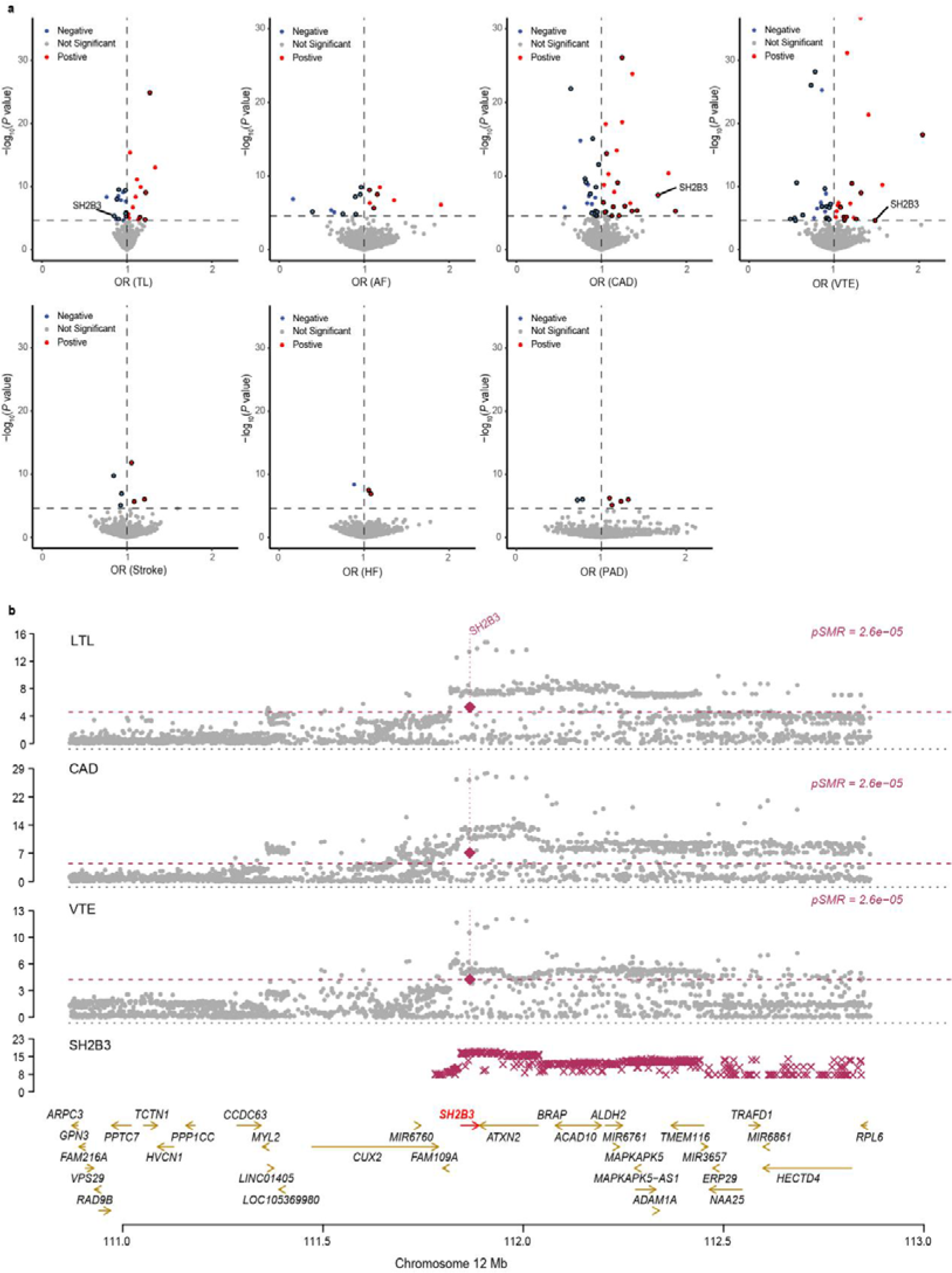
Result summary of Proteome-wide MR analysis on the associations between leukocyte telomere length and six cardiovascular diseases. **(a)** Volcano plots based on SMR showing circulating proteins (red, blue, or gray dots) with the associations between circulating protein levels and each of the traits (x-axis) and the corresponding P-value (y-axis). Red dots indicate positive causal relations. Blue dots indicate negative causal relations. Gray dots indicate insignificant causal relations. Protein-trait associations passing Bonferroni correction (P < 2.60 × 10−5) are outlined in black. **(b)** Miami plots of the 12q24.12 region. Genes that were mapped under the locus at 12q24.12 were highlighted in the analysis. The protein SH2B3 is labeled as significantly associated with LTL, CAD, and VTE. The red horizontal dashed line corresponds to Bonferroni correction (*P* < 2.60 × 10^−5^). LTL, leukocyte telomere length; AF, Atrial fibrillation; CAD, Coronary artery disease; VTE, Venous thromboembolism; HF, Heart failure; PAD, Peripheral artery disease.

## Discussion

In this extensive genome-wide pleiotropy association study, we systematically elucidated the shared genetic architecture beyond genome-wide genetic correlations between LTL and six major CVDs, uncovering no causal relationships between them. Subsequent in-depth analyses identified pleiotropic genetic variants and loci, pleiotropic genes, biological pathways, and protein targets, all reinforcing the involvement of the DNA biosynthesis and telomere maintenance in the shared genetic etiology of these traits. Overall, these findings offer novel insights into the relationship and shared genetic mechanisms underlying LTL and six major CVDs.

Consistent with prior robust epidemiological evidence, our findings reveal weak to moderate, yet significant, negative genome-wide genetic correlations between LTL and six major CVDs, except for LTL-AF. Beyond mere genetic correlations, our analysis uncovers a more extensive degree of genetic overlap between LTL and CVDs using MiXeR and LAVA, involving a broad mixture of both concordant and discordant effect sizes. Employing MiXeR, we demonstrated that the polygenicity of LTL and CVDs presents fundamental distinctions beyond SNP-based heritability. Briefly, LTL was substantially less polygenic than three CVD phenotypes (CAD, HF, and Stroke), yet exhibited similar polygenicity to other CVD phenotypes, including AF, VTE, and PAD. Despite notable differences in polygenicity, we observed extensive genetic overlaps between LTL and all CVDs, supported by LAVA local correlations. This pattern emerged in cases of weak or non-significant genome-wide genetic correlations, such as between LTL and AF, and in strong genome-wide correlations, such as between LTL and PAD. For example, despite the absence of genome-wide genetic correlations, a pronounced fraction of the genetic risk underlying LTL overlaps with AF was indicated using MiXeR. The findings correspond with the discovery of a similar number of positively and negatively correlated genomic regions between LTL and AF by LAVA, alongside further detecting much more shared pleiotropic loci below the genome-wide significance threshold. Although the local genetic correlations encompassed genomic regions averaging approximately 1 megabase (Mb) in width, this resolution required refinement to minimize the impact of heterogeneous effects on estimation accuracy. For example, we noted inconsistent effects across lead variants in the LTL and AF pleiotropic analysis. Specifically, out of 47 top lead variants, 28 demonstrated the same effect direction for LTL and AF, whereas the remaining 19 exhibited opposite effect directions. This indicates that pleiotropic analysis at the level of single variants was necessary to offer further, more detailed insights into the shared genetic underpinnings across complex traits. These findings indicate that genetic overlap between LTL and CVDs was greatly underestimated due to the patterns of mixed effect directions concealed by estimates of genome-wide genetic correlations.

The intricate relationship between LTL and six major CVDs may be further elucidated by examining the vertical and horizontal pleiotropy mechanisms that underpin their shared genetic basis. The causal relationships between LTL and six major CVDs were then predominantly elucidated by the effects attributed to vertical pleiotropy. Using more recent GWAS summary data, we discovered minimal evidence supporting the causal effects of LTL on CVDs and vice versa using LCV and LHC-MR. This finding stood in stark contrast to existing research that suggested such effects were present. Previous two-sample MR studies have reported associations between genetic liability to LTL shortening and the increased risk of CAD and Stroke, but results for the risk of HF have been inconsistent. Another study even demonstrated that genetically predicted AF contributes to the shortening of LTL rather than the reverse. Indeed, the majority of GWAS meta-analyses of CVDs have incorporated samples from the UK Biobank, resulting in considerable sample overlap with GWAS studies on LTL, which might contravene the basic principles of two-sample MR. In addition, the interpretation of the MR estimate in this case was quite complicated by the fact that both LTL and CVDs were time-varying outcomes with a late age of onset. Consequently, our study’s findings suggest that the association between LTL and CVDs may have been greatly overestimated in prior research, potentially as a result of sample overlap, reverse causation, or unmeasured heritable confounding factors, indicating that pleiotropic and common biological pathways may be a better explanation for their association.

The horizontal pleiotropic analyses, encompassing different levels such as pleiotropic genetic variants and loci, pleiotropic genes, biological pathways, and protein targets, revealed significant genetic overlaps between LTL and CVDs from another perspective from another perspective. At the SNP level, pleiotropic variants linking LTL and CVDs were broadly distributed, with a notable emphasis on shared pleiotropic loci among certain trait pairs, including 16q22.1 (*TMED6*), 8p21.3 (*XPO7*), 17p13.3 (*SERPINF1*), and 12q24.12 (*ATXN2*). For example, the transmembrane emp24 domain-containing protein 6 precursor (*TMED6*), highly and selectively expressed in pancreatic islets, was found to be associated with LTL and all CVDs, which belonged to the EMP24_GP25L superfamily and played a crucial role in protein trafficking and secretion. Knockdown of the *TMED6* gene in Min6 β-cells and INS1 cells led to a reduction in glucose-stimulated insulin secretion, suggesting that dysregulation of *TMED6* may play a critical role in the onset of type 2 diabetes^27,28^. To date, there have been no reports of an association between *TMED6* and either LTL or CVDs, hinting at a potential biological mechanism that may be mediated through CVD risk factors, particularly type 2 diabetes. Exportin-7 (*XPO7*) is a bidirectional transporter regulating the nuclear-cytoplasmic shuttling of a wide array of substrates, yet its function remains relatively obscure.^29^. Recent research has identified *XPO7* as a novel regulator of cellular senescence. The depletion of *XPO7* leads to decreased levels of *TCF3* (transcription factor 3, also known as *E2A*) and impaired induction of the cyclin-dependent kinase inhibitor p21^CIP1^, which was critical during oncogene-induced senescence^30^. The role and malfunction of the Serpin Family F Member 1 (*SERPINF1*), also known as the pigment epithelium-derived factor (*PEDF*) gene, in the aging process has currently been a hot topic. Briefly, the reduction in *PEDF* expression levels, both directly and indirectly, can prompt cellular senescence via the modulation of various signaling pathways^31,32^. Studies have also demonstrated that *PEDF* possesses insulin-sensitizing effects in the liver and adipose tissues and exhibits anti-inflammatory, anti-thrombogenic, and vasculoprotective properties in vivo, offering protection against metabolic syndrome and cardiovascular diseases^33,34^. Overall, this study extends previous findings on shared genetic architecture by offering a more comprehensive characterization of specific pleiotropic loci.

At the gene level, we employed two gene-mapping strategies to identify credible mapped genes for all jointly associated pleiotropic loci. Seven genes, namely *ALDH2*, *ACAD10*, *TMEM116*, *SH2B3* (all located at 12q24.12), *TMED6* (16q22.1), *SERPINF1* (17p13.3), and *XPO7* (8p21.3), have emerged as the most pleiotropic; they are thought to have important regulatory functions, influencing over half of the trait pairs examined. For example, among these genes, *ALDH2* and *ACAD10*, located at the 12q24.12 locus, were associated with LTL and all CVDs except AF. Aldehyde dehydrogenase 2 (*ALDH2*) is the gene with the highest number of genetic polymorphisms in humans; it is involved in encoding a mitochondrial enzyme critical for detoxifying reactive aldehydes. For example, the *ALDH2* rs671 inactivating polymorphism, found in up to 8% of the global population and up to 50% of the East Asian population, is associated with an elevated risk of several CVDs, such as CAD. While numerous studies have connected aldehyde accumulation, due to alcohol consumption, ischemia, or heightened oxidative stress, to elevated CVD risk, this accumulation alone does not fully account for their complex interactions^35,36^. Moreover, previous studies indicated that the *ALDH2* enzyme might also exacerbate myocardial remodeling and contractile dysfunction during aging, potentially via AMPK/Sirt1-mediated mitochondrial damage. Acyl-CoA dehydrogenase family member 10 (*ACAD10*) gene encoded an enzyme involved in fatty acid beta-oxidation in mitochondria^37^. *ACAD10* is predominantly expressed in the human brain and is believed to play a role in physiological functions within the central nervous system, such as the regulation of cellular lipid synthesis. *ACAD10* has previously been identified as one of the putative causal genes for CAD, stroke, and hypertension, a common risk factor for CVDs^38^. Collectively, these findings imply that *ACAD10* has a significant role in regulating lipid synthesis through distinct molecular and cellular pathways, potentially influencing the development of CVDs. Further research is required to elucidate the precise mechanisms of interaction between ACAD10 and both LTL or CVDs.

The 12q24.12 locus was a top hit region, which identified as pleiotropic for all correlated trait pairs except for LTL-AF and exhibited strong evidence of colocalization between these trait pairs. Besides, HyPrColoc further revealed robust colocalization evidence for this locus between LTL and all CVDs, excluding AF and PAD, encompassing the shared causal SNP (i.e., rs10774625). LAVA results supported these findings and showed consistent negative local genetic correlations between LTL and all CVDs, except for AF, HF and PAD. The index SNP rs10774625 polymorphism has been reported to be associated with a variety of CVDs, notably CAD, alongside cardiometabolic markers such as blood pressure and blood lipids. The index SNP rs10774625, located at the 12q24.12 locus (an intronic variant of the *ATXN2* gene), was associated with eQTLs and pQTLs in whole blood for the SH2B adaptor protein 3 (*SH2B3*). The proteome-wide Mendelian Randomization study revealed that the genetically predicted levels of SH2B3 protein were significantly associated with both LTL-CAD and LTL-VTE. *SH2B3* is a member of the adapter protein family and plays a critical role in negatively regulating cytokine signaling and cell proliferation. It was originally described as a regulator of hematopoietic and lymphocyte differentiation and was implicated in the transduction and regulation of growth factors and inflammation-related cytokine receptor-mediated signaling. *SH2B3* missense variants may affect lifespan through cardiovascular disease^39^. Specifically, *SH2B3* can lead to increased production of IFNγ, which acts as a pro-inflammatory mediator to induce the polarization of macrophages into different states. This process is crucial for mediating inflammatory regulation and fibrosis post-myocardial infarction^40^. On the other hand, IFNγ is released and activated by CD4 T helper cells—specifically Th1 cells—which are instrumental in coordinating immune cell infiltration and inflammation. The above immune cells and inflammatory signals are pivotal in the progression of non-ischemic heart failure in patients^41^. Notably, increased cardiomyocyte size and fibrosis are critical characteristics of cardiac hypertrophy and remodeling, which ultimately lead to heart failure^42^. Further studies have demonstrated that cardiac-specific *SH2B3* overexpression exacerbates pressure overload, leading to cardiac hypertrophy, fibrosis, and dysfunction by activating focal adhesion kinase, which subsequently triggers the downstream phosphoinositide 3-kinase-AKT-target of Programmed Death-1 (PD-1). Significant overexpression of the *SH2B3* gene promotes the activation of the Akt signaling pathway, which can promote cardiac hypertrophy and fibrosis and lead to the deterioration of cardiac function^43^. Meanwhile, age-related telomere dysfunction is a core driver of inflammation^44^. Therefore, *SH2B3* may become one of the most promising therapeutic targets for CVDs. In contrast, *TMEM116*, a member of the TMEM family of proteins that spans the plasma membrane of cells to facilitate intercellular communication, demonstrates a different aspect of disease association. Multiple members of the TMEM family may be up or down-regulated in tumor tissues, and some of them are used as cancer prognostic biomarkers^45^. However, in studies of cardiovascular diseases, this protein has only been found to be related to coronary atherosclerosis^46^, and the specific mechanism is unclear and requires further research.

At the pathway level, functional analyses of the pleiotropic loci between LTL and CVDs have implicated genes involved in the metabolic processes of nucleobase-containing compounds, including DNA biosynthesis and telomere maintenance. There is a close relationship between telomere maintenance, telomerase expression, and extension of cell lifespan. Telomeres undergo shortening during repeated cell divisions, and when their length diminishes to a critical point, the resultant genomic instability can lead to further genetic abnormalities that promote cell death or apoptosis, which is a hallmark of cellular senescence. Estrogen, stress accumulation from oxidative damage, hypertension, etc., are believed to significantly impact telomere homeostasis and contribute to the development of CVDs^47^. This finding bolsters the theory that progressive telomere shortening contributes to the pathogenesis of age-related human diseases such as CVDs. Specifically, Minano et al. documented the presence of vascular endothelial cells exhibiting age-related phenotypes within human atherosclerotic lesions^48^. Excessive vascular smooth muscle cells are stimulated to proliferate and migrate, leading to the growth of atherosclerosis. Besides, telomerase activation and telomere maintenance are critical in increasing the proliferation and growth of vascular smooth muscle cells. Activation through the telomerase reverse transcriptase component (TERT) extends the lifespan of cultured vascular smooth muscle cells. Conversely, telomerase inhibition can extend the lifespan and reduce the proliferation of cultured vascular smooth muscle cells, thereby decreasing the risk of atherosclerosis^49^. Therefore, these findings highlight the role of telomeres in cardiovascular health, suggesting that methods to modulate the balance of telomerase activity may be critical for developing effective interventions for related CVDs.

There were some limitations to the current study. Firstly, the main analysis focused solely on individuals of European ancestry due to the scarcity of sufficiently powered GWAS involving other ancestries. Nevertheless, we utilized GWAS summary data from East Asian ancestries for replication, partially confirming the consistency of the genetic foundation identified in the European sample. Future studies with a trans-ancestral approach are necessary to evaluate the universality of these findings. Secondly, the analysis focused on common genetic variants that account for only a small fraction of overall disease risk. The remaining variance was likely attributable to many undetected SNPs, rare variants, or gene interactions. Therefore, they need to be further studied to achieve a more complete understanding. Thirdly, although we uncovered the potential shared genetic architecture, the mechanisms of shared biological pathways still require further experimental validation. Finally, our analysis of GWAS summary data encompassed six major CVDs, representing a significant portion of the genetic risk architecture for these conditions, though not comprehensively. As GWAS datasets expand, it will be crucial to undertake cross-trait analyses incorporating more varied datasets and additional diseases to enhance our understanding.

## Conclusion

In conclusion, we found extensive polygenic overlap between LTL and CVDs, with distinct patterns of genetic correlations and effect directions, uncovering no causal relationships. It was proved that LTL and six major CVDs share pleiotropic genetic variants, loci, genes, biological pathways, and protein targets, underscoring the role of DNA biosynthesis and telomere maintenance in their common genetic etiology. These findings elucidate the interconnected mechanisms between LTL and CVDs, potentially guiding targeted therapies and clinical practice.

## Methods

### Data Sources and Quality Control

Figure 1 outlines the workflow for our study. Due to the confounding effects of ancestral differences in linkage disequilibrium (LD) structure and the scarcity of sufficiently large multi-ancestry samples, we limited our main analysis to individuals of European ancestry. We sourced genome-wide association study (GWAS) summary statistics from the most comprehensive and recent publicly available datasets of European ancestry. Specifically, GWAS summary statistics for leukocyte telomere length (LTL) were derived from a published GWAS comprising 464,716 individuals of European ancestry from the UK Biobank^7^. LTL was quantified as the ratio of telomere repeats copy number (T) to a single copy gene (S) in a mixed leukocyte population, measured via a multiplex quantitative polymerase chain reaction (qPCR) assay, and subsequently log-transformed to achieve an approximation to a normal distribution. Our selection criteria for GWAS included studies with sample sizes exceeding 50,000 to ensure adequate statistical power. Accordingly, we included GWAS summary statistics for six major cardiovascular diseases (CVDs): atrial fibrillation (AF)^8^, coronary artery disease (CAD)^9^, venous thromboembolism (VTE)^10^, heart failure (HF)^11^, peripheral artery disease (PAD)^12^, and stroke^13^. AF GWAS summary statistics were sourced from a genome-wide meta-analysis of six studies (The Nord-Trøndelag Health Study [HUNT], deCODE, the Michigan Genomics Initiative [MGI], DiscovEHR, UK Biobank, and the Atrial Fibrillation Genetics [AFGen] Consortium), encompassing 60,620 AF cases and 970,216 controls of European ancestry. For CAD, we utilized GWAS summary statistics from a genome-wide meta-analysis by the CARDIoGRAMplusC4D Consortium and the UK Biobank, which included 181,522 cases and 984,168 controls. VTE GWAS summary statistics were extracted from a meta-analysis of 81,190 cases and 1,419,671 controls of European ancestry across 7 cohorts (the Copenhagen Hospital Biobank Cardiovascular Disease Cohort [CHB-CVDC], Danish Blood Donor Study [DBDS], deCODE, Intermountain Healthcare, UK Biobank, FinnGen, and Million Veterans Program [MVP] Consortium). GWAS summary statistics for HF came from the Heart Failure Molecular Epidemiology for Therapeutic Targets (HERMES) Consortium, including 47,309 cases and 930,014 controls. GWAS summary statistics for PAD were derived from a genome-wide meta-analysis of 11 independent GWASs, totaling 12,086 cases and 499,548 controls. Lastly, GWAS summary statistics for Stroke were obtained from the GIGASTROKE consortium, which comprised 73,652 cases and 1,234,808 controls of European ancestry. Detailed information about these GWAS summary statistics and their original publication sources is available in Supplementary Table 1.

Prior to further analysis, stringent quality control measures were applied to the GWAS summary statistics, encompassing several key steps: (i) alignment with the hg19 genome build, referencing the 1000 Genomes Project Phase 3 Europeans; (ii) restriction of the analysis to autosomal chromosomes; (iii) removal of single nucleotide polymorphisms (SNPs) lacking a rsID or presenting duplicated rsIDs; and (iv) exclusion of rare or low-frequency variants, defined by a minor allele frequency (MAF) less than 1%. To ensure robust and interpretable comparisons between LTL and CVDs, we standardized the summary statistics to include only SNPs present across all analyzed phenotypes, resulting in a cohesive dataset of 6,923,146 SNPs. Additionally, in subsequent analyses, we implemented further data processing techniques tailored to the specific requirements of various statistical tools.

### Genetic overlap

To explore the shared genetic foundations between LTL and six major CVDs, we evaluated genetic overlap across genome-wide, polygenic, and local levels.

### Genome-wide genetic correlation analysis between LTL and CVDs

At the genome-wide level, we analyzed SNP-level heritability for each trait and the genetic correlations (*r_g_*) between LTL and six major CVDs using cross-trait linkage disequilibrium (LD) score regression (LDSC)^50,51^. LDSC facilitates the estimation of the average genetic effect sharing across the entire genome between two traits, leveraging GWAS summary statistics. This includes the contribution of SNPs below the threshold of genome-wide significance and accounts for potential confounding factors such as polygenicity, sample overlap, and population stratification. First, SNP-based heritability (*h^2^_SNP_*, representing the fraction of phenotypic variation explained by common genetic variations included in the study) for LTL and six major CVDs was estimated using univariate LDSC. This analysis utilized pre-computed LD scores from the European reference panel in the 1000 Genomes Project Phase 3, excluding SNPs that did not overlap with the reference panel. Notably, the major histocompatibility complex (MHC) region (chr 6: 25-35 Mb), known for its intricate LD structure, was omitted from the main analysis. Second, bivariate LDSC analysis estimated the genetic correlations between LTL and the six major CVDs. This method utilizes a weighted linear model, where it regresses the product of Z-statistics from two traits against the LD score across all genetic variants genome-wide. Genetic correlations with *P*-values below the Bonferroni-adjusted threshold (*P* = 0.05 / number of trait pairs = 0.05 / 6 = 8.33×10^−3^) were deemed statistically significant.

To elucidate the biological underpinnings of the shared genetic predisposition to LTL and six major CVDs, we employed stratified LDSC applied to specifically expressed genes (LDSC-SEG) to identify relevant tissue and cell types. This analysis incorporated tissue and cell type-specific expression data from the Genotype-Tissue Expression (GTEx) project and the Franke lab, covering 53 tissues and 152 cell types. Additionally, we utilized chromatin-based annotations associated with six epigenetic marks (DNase hypersensitivity, H3K27ac, H3K4me1, H3K4me3, H3K9ac, and H3K36me3) for validation purposes. These annotations included 93 labels from the Encyclopedia of DNA Elements (ENCODE) project and 396 labels from the Roadmap Epigenomics database. For the identified relevant tissues or cell types, we adjusted the *P*-values for the significance of the coefficients using the False Discovery Rate (FDR) method. An FDR threshold of < 0.05 was established as the criterion for statistical significance.

### Polygenic overlap analysis between LTL and CVDs

To augment the genome-wide genetic correlation analysis, we engaged the causal mixture modeling approach (MiXeR) to quantify the polygenic overlap between LTL and six major CVDs, independent of genetic correlation directions. MiXeR estimates the quantity of shared and phenotype-specific “causal” variants that exert non-zero additive genetic effects, accounting for 90% of SNP-heritability in each trait^52^. This 90% SNP-heritability threshold minimizes the influence of variants with negligible effects. Importantly, MiXeR’s capacity to evaluate polygenic overlap without regard to the directional effects of variants offers a nuanced view of local genetic associations that might be obscured in traditional genome-wide genetic correlation estimations due to opposing variant effects. Initially, univariate MiXeR analyses estimated the “causal” variant count for LTL and six major CVDs, assessing both polygenicity and the average magnitude of additive genetic effects among these variants. The LD structure was determined using the genotype reference panel from the 1000 Genomes Project Phase 3. The MHC region (chr 6: 25-35 Mb), known for its intricate LD structure, was excluded from the main analysis. Subsequently, bivariate MiXeR analysis quantified the polygenic overlap between LTL and the six CVDs. This analysis delineated the additive genetic effects across four categories: (i) SNPs with zero effect on both traits, (ii) trait-specific SNPs with non-zero effects on the first trait, (iii) trait-specific SNPs with non-zero effects on the second trait, and (iv) SNPs affecting both traits. The Dice coefficient (DC), representing the proportion of shared SNPs between two traits relative to the total number of SNPs associated with either trait, was used to estimate polygenic overlap. Additionally, MiXeR calculated the overall genetic correlations (*r_g_*), the correlation of effect sizes within the shared genetic component (*r_g_s*), and the fraction of variants with concordant effects in the shared component. The model’s predictive accuracy was evaluated through comparison of the modeled versus actual data, utilizing conditional quantile-quantile (Q-Q) plots, log-likelihood plots, and the Akaike information criterion (AIC). Positive AIC differences are interpreted as evidence that the best-fitting MiXeR estimates are distinguishable from the reference model. A negative AIC value indicates that the MiXeR model fails to distinguish effectively between maximal and minimal overlap scenarios.

### Local genetic correlation between LTL and CVDs

To investigate whether there are any genomic loci with pronounced genetic correlations despite negligible genome-wide *r_g,_* we utilized the Local Analysis of [co]Variant Annotation (LAVA) method to estimate the localized genetic correlation between LTL and six major CVDs. LAVA, grounded in a fixed-effects statistical model, enables the estimation of local SNP heritability (loc-*h^2^_SNP_*) and genetic correlations (loc-*r_g_s*) across 2,495 semi-independent genetic regions of approximately equal size (∼1 Mb). This method is adept at identifying loci with mixed effect directions, offering a nuanced measure of genome-wide genetic overlap, albeit influenced by the statistical power of the underlying GWAS data. For this analysis, we adopted the genomic regions defined by Werme et al. as autosomal LD blocks, characterized by minimal inter-LD block linkage with an average size of 1 million bases, each containing at least 2500 variants. The LD reference panel from the 1000 Genomes Project Phase 3 for European samples was employed, and consistent with LDSC and MiXeR analyses, the MHC region (chr 6: 25-35 Mb) was excluded. Identifying meaningful loc-*r_g_s* requires a significant local genetic signal; thus, a stringent *P*-value threshold (*P* < 1×10^−4^) was applied to filter out non-significant loci. Subsequent bivariate testing on selected loci and traits with notable univariate genetic signals was conducted. For these loci, P-values for loc-*r_g_s* were adjusted using the Benjamini-Hochberg FDR method, with an FDR < 0.05 establishing statistical significance. LAVA incorporated an estimate of sample overlap (genetic covariance intercept) from bivariate LDSC analyses to account for potential sample overlap effects.

In cases where shared risk loci were identified across multiple phenotypes, the Hypothesis Prioritisation for multi-trait Colocalization (HyPrColoc) method was employed to assess whether association signals across more than a pair of traits were colocalized. HyPrColoc, an efficient deterministic Bayesian clustering algorithm, leverages GWAS summary statistics to identify clusters of colocalized traits and potential causal variants within a genomic locus, providing a posterior probability (PP) of colocalization for each cluster. Loci with a PP > 0.7 were deemed colocalized, enhancing our understanding of shared genetic architectures across traits.

### Pleiotropy insights to dissect genetic overlap

#### Causal inference between LTL and CVDs

To elucidate the potential causal relationships underlying the genetic correlations observed between LTL and six major CVDs, we employed latent causal variable (LCV) analysis. This method posits that the genetic correlation between two traits operates through a latent factor, allowing for the distinction between genetic causality (vertical pleiotropy) and both correlated and uncorrelated horizontal pleiotropy^53^. It achieves this by estimating the genetic causality proportion (GCP) across all genetic variants, where GCP quantifies the share of each trait’s heritability explained by a mutual latent factor. It is essential to emphasize that GCP does not indicate the magnitude of causal effects but only implies a causal relationship between traits. This method offers insights to evaluate whether the impact of one trait on the second exceeds the evidence in the reverse direction. The sign of genetic correlation can be employed to infer the consequence of the partial genetic causality of one trait on another. A GCP greater than zero suggests a partial genetic causal relationship from trait 1 to trait 2 and vice versa, with values closer to |GCP| = 1 indicating stronger evidence of vertical pleiotropy. Conversely, a GCP of zero implies horizontal pleiotropy. We considered an absolute GCP estimate (|GCP|) greater than 0.60 as indicative of substantial genetic causality, applying a Bonferroni-corrected significance threshold of *P* < 8.33×10^−3^ to account for multiple comparisons across trait pairs. The LCV analysis, while robust, assumes a singular, unidirectional latent variable driving the genetic correlation, a premise potentially confounded by bidirectional causal effects or multiple latent factors.

Addressing the limitations inherent to LCV, we also conducted an analysis using Latent Heritable Confounder Mendelian Randomization (LHC-MR)^54^. This advanced Mendelian randomization approach leverages genome-wide variants to explore bi-directional causal associations between complex traits, thus enhancing the capacity to discern bi-directional genetic effects, direct heritabilities, and confounder effects, even amidst sample overlap. By modeling an unobserved heritable confounder affecting both exposure and outcome traits, LHC-MR assures the critical assumption of exchangeability^55^. Its comprehensive modeling of potential SNP effects-direct, indirect, or null-on the traits offers more accurate causal effect estimates compared to traditional MR methods, such as MR Egger, weighted median, inverse variance weighted (IVW), simple mode, and weighted mode^56–58^. For statistical significance, we adjusted for multiple testing with a threshold of *P* < 4.17×10^−3^, considering both the number of trait pairs and the number of tests conducted.

#### Pairwise Pleiotropic analysis between LTL and CVDs

To delve into the role of horizontal pleiotropy between LTL and six major CVDs, we utilized the Pleiotropic Analysis under the Composite Null Hypothesis (PLACO) method to conduct a comprehensive genome-wide identification of pleiotropic SNPs that concurrently influence the risk of both traits simultaneously. PLACO operates on the composite null hypothesis that asserts a genetic variant is either associated with just one or neither trait, thereby distinguishing between pleiotropic effects and singular trait associations^59^. The method evaluates this hypothesis by examining the product of the Z statistics derived from the GWAS summary statistics of both traits, formulating a null distribution of the test statistic as a mixture distribution. This allows for the acknowledgment of SNPs that may be linked to only one or none of the phenotypes under study. The threshold for identifying pleiotropic SNPs with significant evidence of genome-wide pleiotropy was set at *P_PLACO_* < 5×10^−8^. Additionally, to adjust for possible sample overlap, we de-correlated the Z-scores using a correlation matrix directly estimated from the GWAS summary statistics, ensuring a more accurate interpretation of pleiotropic effects.

### Characterization of pleiotropic loci and functional annotation

The Functional Mapping and Annotation of Genome-Wide Association Studies (FUMA) platform was employed to identify independent genomic loci and conduct functional annotation for pleiotropic SNPs revealed by PLACO analysis^59^. FUMA, leveraging data from 18 biological databases and analytical tools, annotates GWAS findings to highlight probable causal genes through positional and eQTL mapping ^60^. The 1000 Genomes Project Phase 3 European-based LD reference panels were utilized for LD structure correction. Initially, FUMA distinguishes independent significant SNPs (meeting genome-wide significance at *P* < 5×10^−8^ and r^2^ < 0.6), further defining a subset as lead SNPs based on mutual independence (r^2^ < 0.1). LD blocks within 500 kb of lead SNPs are merged to delineate distinct genomic loci, with the SNP exhibiting the lowest *P*-value in each locus designated as the top lead SNP. The analysis then assesses directional effects between LTL and six major CVDs by comparing Z-scores of these top lead SNPs. SNPs achieving genome-wide significance (*P* < 5×10^−8^) in individual GWAS for each trait were annotated using FUMA for comparative analysis. The identified pleiotropic loci were considered novel if they did not coincide with the loci previously reported in the original GWAS for LTL or any of the six major CVDs. In other words, to be deemed ‘novel,’ a locus identified through FUMA should not have exhibited statistical significance in the single trait GWAS.

To elucidate the biological underpinnings of the observed statistical associations, lead SNPs were annotated using Annotate Variation (ANNOVAR)^61^ for their proximity to genes and potential impact on gene function. The Combined Annotation-Dependent Depletion (CADD) score^62^, which aggregates insights from 67 annotation resources, was used to assess the deleteriousness of variants. Variants with CADD scores greater than 12.37 were deemed likely to exert deleterious effects. Furthermore, the RegulomeDB score provided a categorical assessment of an SNP’s regulatory potential based on expression quantitative trait loci (eQTL) and chromatin marks, ranging from 1 (strong evidence of regulatory functionality) to 7 (minimal evidence)^63^. The highest regulatory potential is indicated by a score of 1a, while a score of 7 suggests the least regulatory significance. Chromatin states, determined by ChromHMM using data from 127 epigenomes and five chromatin marks, revealed the genomic regions’ accessibility, categorized into 15 states. For the identification of putative causal genes, SNPs were mapped using two approaches: positional mapping within a 10-kb window around the SNP and eQTL mapping.

### Colocalization analysis

For pleiotropic loci identified and annotated by FUMA, we conducted a colocalization analysis using COLOC to pinpoint potential shared causal variants across pairwise traits within each locus. COLOC evaluates five mutually exclusive hypotheses for each pair of traits at a locus^64^: H0 posits no association with either trait; H1 and H2 suggest an association with only one of the traits; H3 indicates that both traits are associated due to different causal variants; and H4 implies a shared association for both traits stemming from the same causal variant. The analysis was performed using default COLOC prior probabilities: p1 and p2, each set at 1×10^−4^ for an SNP’s association with the first and second trait, respectively, and p12 at 1×10^−5^ for an SNP associated with both traits. A Posterior Probability for Hypothesis 4 (PP.H4) greater than 0.7 was considered strong evidence for colocalization, suggesting the presence of shared causal variants at the locus. The SNP exhibiting the highest PP.H4 was identified as a candidate causal variant.

### Gene level analyses

Building on the insights from PLACO, we delved into the shared biological processes and pathways involving the identified pleiotropic loci. Through gene-level analysis using Multi-marker Analysis of GenoMic Annotation (MAGMA), we assessed genes within or intersecting the pleiotropic loci, integrating data from both PLACO and single-trait GWAS. Unlike permutation-based approaches, MAGMA employs a multiple regression model that incorporates principal component analysis to evaluate gene associations. This model calculates a p-value for each gene, aggregating the impact of all SNPs linked to that gene while considering gene size, SNP count per gene, and linkage disequilibrium (LD) among the markers. SNPs were attributed to genes based on their location within the gene body or within a 10 kb range upstream or downstream. The LD calculations leveraged the 1000 Genomes Project Phase 3 European population as the reference panel, with SNP locations determined using the human genome Build 37 (GRCh37/hg19) and focusing on 17,636 autosomal protein-coding genes. A gene was deemed significant if its p-value was below 0.05 after applying a Bonferroni correction for the total number of protein-coding genes and the six trait pairs analyzed (*P* = 0.05 / 17,636 / 6 = 4.73×10^−7^). Due to complex LD patterns, the MHC region (chr6: 25-35 Mb) was excluded from MAGMA’s gene-based analysis.

To overcome the limitations of MAGMA, which assigns SNPs to their nearest genes based on arbitrary genomic windows potentially missing functional gene associations due to long-range regulatory effects, we employed eQTL-informed MAGMA (e-MAGMA) for a more nuanced investigation of tissue-specific gene involvement based on PLACO results. e-MAGMA retains the statistical framework of MAGMA, using a multiple linear principal component regression model, but enhances gene-based association analysis by incorporating tissue-specific cis-eQTL information for SNP assignment to genes, which yields more biologically relevant and interpretable findings. For our analysis, we utilized eQTL data from 47 tissues provided by the GTEx v8 reference panel, as available on the e-MAGMA website. Guided by the principle that analyses focused on disease-relevant tissues yield more pertinent insights, we selected ten relevant tissues for our study. These included three artery tissues, two adipose tissues, two heart tissues, and three additional tissues (whole blood, liver, and EBV-transformed lymphocytes), chosen based on their significant enrichment in the LDSC-SEG analysis. The LD reference data for our analysis came from the 1000 Genomes Phase 3 European panel. We calculated tissue-specific p-values for each gene across the selected tissues, with significance determined post-Bonferroni correction for the number of tissue-specific protein-coding genes and trait pairs examined. For instance, the significance threshold for adipose subcutaneous tissue was set at *P* = 0.05 / 9,613 / 6 = 8.67×10^−7^. Similar to MAGMA, e-MAGMA analysis results within the MHC region (chr6: 25-35 Mb) were excluded to avoid confounding due to complex LD patterns. Additionally, we conducted a transcriptome-wide association study (TWAS) based on single-trait GWAS results using the functional summary-based imputation software, FUSION, applying tissue-specific Bonferroni corrections to determine significance. The FUSION approach integrates GWAS summary statistics with pre-computed gene expression weights, referencing the same tissues analyzed in the e-MAGMA study from the GTEx v8 dataset. This integration takes into account the LD structures to identify significant relationships between gene expression levels and specific traits.

### Pathway level analyses

To investigate the genetic pathways underlying the comorbidity of LTL and six major CVDs, we utilized MAGMA for gene-set analysis. This analysis employs a competitive approach, where test statistics for all genes within a gene set, such as a biological pathway, are aggregated to derive a joint association statistic. Gene sets were sourced from the Gene Ontology biological processes (GO_BP) via the Molecular Signatures Database (MsigDB), with gene definitions and association signals derived from MAGMA gene-based analysis. We adjusted for multiple testing using a Bonferroni correction, setting the threshold at *P* = 0.05 / 7,744 / 6 = 1.08×10^−6^. We then conducted functional enrichment analysis on the genes overlapping across more than one trait pair, which were significantly identified by both MAGMA and e-MAGMA analyses. For this purpose, the ToppGene Functional Annotation tool (ToppFun) was employed to identify significantly represented biological processes and enriched signaling pathways, considering the entire genome as the background. ToppFun performs Functional Enrichment Analysis (FEA) on the specified gene list, leveraging a broad spectrum of data sources, including transcriptomics, proteomics, regulomics, ontologies, phenotypes, pharmacogenomics, and bibliographic data. The list of candidate genes was submitted to the ToppFun tool within the ToppGene Suite, with an FDR < 0.05 established as the threshold for statistical significance.

### Proteome-wide Mendelian Randomization study analysis

To explore potential common causal factors at the proteomic level, we utilized Summary data-based Mendelian Randomization (SMR) to examine associations between protein abundance and disease phenotype. This analysis leveraged index cis-acting variants (cis-pQTLs) identified in the UK Biobank Pharma Proteomics Project (UKB-PPP), which includes plasma samples from 34,557 European individuals with the measurement of 2,940 plasma proteins using the Olink Explore platform. Cis-pQTLs were defined as SNPs located within a 1Mb radius from the transcription start site (TSS) of the gene encoding the protein. Only index cis-pQTLs associated with plasma protein levels at a genome-wide significance threshold (*P* < 5×10^−8^) were considered for inclusion in the SMR analysis. Summary data-based Mendelian Randomization (SMR) is designed to prioritize genes for which expression levels are potentially causally linked to an outcome trait, utilizing summary statistics within a Mendelian Randomization framework. To differentiate between pleiotropy and linkage (where protein abundance and a phenotype manifestation could be influenced by two separate causal variants in strong linkage disequilibrium with one another), the Heterogeneity in Dependent Instrument (HEIDI) test was employed. A HEIDI test *p*-value below 0.01 signifies the presence of two distinct genetic variants in high linkage disequilibrium, explaining the observed associations. Additionally, to address potential biases from analyzing single SNPs, a multi-SNP approach (multi-SNPs-SMR) was utilized as a sensitivity analysis, enhancing the reliability of the statistical evidence. A p-value less than 0.05 in the multi-SNPs-SMR analysis was deemed significant. Furthermore, HyPrColoc analysis was employed to ascertain whether the associations identified between proteins and various diseases stemmed from the same causal variant or were due to linkage disequilibrium. A posterior probability of a shared causal variant (PP.H4) greater than 0.7 signifies strong evidence of colocalization between proteins and multiple diseases.

## Reporting summary

Further information on research design is available in the Nature Portfolio Reporting Summary linked to this article.

## Data availability

The study used only openly available GWAS summary statistics on leukocyte telomere length and six major cardiovascular diseases that have originally been conducted using human data. GWAS summary statistics on LTL are available at https://figshare.com/s/caa99dc0f76d62990195. GWAS summary statistics on AF, HF, and Stroke are available at the GWAS Catalog (GCST90104539, GCST009541, and GCST90104539). GWAS summary statistics on CAD and PAD are publicly available for download at the Cardiovascular Disease Knowledge Portal (CVDKP) website: https://cvd.hugeamp.org/datasets.html. GWAS summary statistics on VTE are obtained from the deCODE genetics website: https://www.decode.com/summarydata/. Blood-based cis-pQTL from UKB-PPP are obtained from https://www.synapse.org/#!Synapse:syn51365303, respectively.

## Code availability

All software used to conduct the analyses in this paper are freely available online. Software (version, where applicable) and sources are listed below: LDSC (v1.0.1; https://github.com/bulik/ldsc), MiXeR (v1.3; https://github.com/precimed/mixer), LAVA (v0.1.0; https://github.com/josefin-werme/LAVA), LCV (https://github.com/lukejoconnor/ LCV); LHC-MR (v0.0.0.9000; https://github.com/LizaDarrous/lhcMR), PLACO (v0.1.1; https://github.com/RayDebashree/PLACO), FUMA (v1.5.4; http://fuma.ctglab.nl/), HyPrColoc(v1.0; https://github.com/jrs95/hyprcoloc), MAGMA (v.1.08; https://ctg.cncr.nl/ software/magma), e-MAGMA (https://github.com/eskederks/eMAGMA-tutorial), TWAS (http://gusevlab.org/projects/fusion/), SMR (v1.31; https://yanglab.westlake.edu.cn/ software/smr/), COLOC (v5.2.1; https://github.com/chr1swallace/coloc), and R (v.4.1.3; https://www.r-project.org/).

## Acknowledgements

This study was supported by the Natural Science Foundation of China Excellent Young Scientists Fund (Overseas) (Grant no. K241141101), Guangdong Basic and Applied Basic Research Foundation for Distinguished Young Scholars (Grant no. 24050000763), Shenzhen Pengcheng Peacock Plan, Shenzhen Basic Research General Projects of Shenzhen Science and Technology Innovation Commission (Grant no. JCYJ20230807093514029) (To Y.F.), National Natural Science Foundation (Grant no. 82170339 and 82270241), NSFC Incubation Project of Guangdong Provincial People’s Hospital (Grant no. KY0120220021), Natural Science Foundation of Guangdong Province (Grant no. 2023B1515020082) (To L.J.), National Natural Science Foundation of China (Grant no. 82260073); Tianshan Talent Cultivation Program Project of Xinjiang Uygur Autonomous Region (Grant no. 2022TSYCLJ0028) (To Y.Y.), and Center for Computational Science and Engineering at Southern University of Science and Technology. The funder had no role in the design, implementation, analysis, interpretation of the data, approval of the manuscript, and decision to submit the manuscript for publication.

## Author contributions

J.Q., Y. F., Y.Y., L.J., and S.P. conceptualized and supervised this project and wrote the manuscript. J.Q., Q.W., and Y.Z. performed the main analyses and wrote the manuscript. J.Q., M.C., L.C., and F.L. performed the statistical analysis and assisted with interpreting the results. K.Y., L.Z., N.T., P.H., and A.J. provided expertise in cardiovascular biology and GWAS summary statistics. All authors discussed the results and commented on the paper.

