## Supplementary Figures and Notes for "Contribution of leukocyte telomere length to major cardiovascular diseases onset: phenotypic and genetic insights from a large-scale genome-wide cross-trait analysis"

Professor

Department of Cardiology, People's Hospital of Xinjiang Uygur Autonomous Region, Urumqi, , Xinjiang Uygur Autonomous Region, 830000, China.

Xinjiang Key Laboratory of Cardiovascular Homeostasis and Regeneration Research, Urumqi, Xinjiang Uygur Autonomous Region, 830001, China.

Lei Jiang MD, PhD

Associate Professor

Department of Cardiology; Guangdong Cardiovascular Institute, Guangdong Provincial People's Hospital (Guangdong Academy of Medical Sciences); Southern Medical University, Guangzhou, Guangdong, 510080, China.

Siim Pauklin PhD

Group leader and CRUK Career Development Fellow

Botnar Research Centre, Nuffield Department of Orthopaedics, Rheumatology and Musculoskeletal Sciences, University of Oxford, Headington, Oxford OX3 7LD, UK.

**Abstract**

Telomere shortening, a marker of cellular aging and genomic instability, has been epidemiologically linked to an increased risk of various cardiovascular diseases (CVDs). However, shared genetic determinants involved in these associations remain unclear. We composed an atlas of the shared genetic associations between leukocyte telomere length (LTL) and six major CVDs by investigating shared genetic elements, encompassing SNPs, genes, biological pathways, and protein targets with pleiotropic implications. Extensive genetic overlaps beyond genetic correlations were observed, but no causal relationships were established. We identified 248 independent pleiotropic genomic risk loci, implicating 50 unique genes in two or more trait pairs, especially the *SH2B3* gene, which was further validated by a proteome-wide Mendelian Randomization study. Functional analysis demonstrated a link to both DNA biosynthetic processes and telomere maintenance mechanisms. These findings suggest a genetic link between LTL and CVDs, highlighting a shared genetic basis crucial for developing future interventions and therapeutic targets.

**Supplementary Figures and Notes**

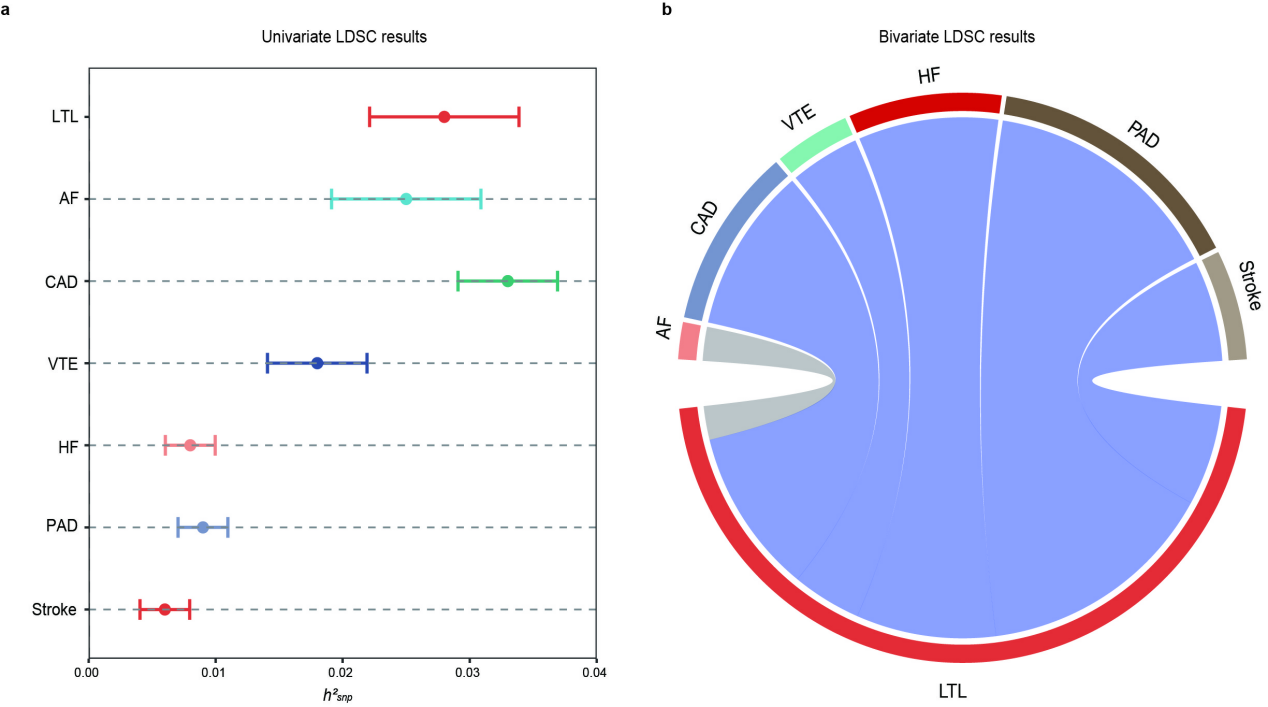
**Supplementary Fig. 1. Supplemental LDSC figures for leukocyte telomere length and six cardiovascular diseases.**

**(a)** Error-bar plot of the SNP-based heritability (*h^2^_SNP_*) point estimates for leukocyte telomere length and six major cardiovascular diseases, computed by univariate LDSC. **(b)** Network visualization of the Bonferroni-corrected significant global *r_g_* between leukocyte telomere length and six major cardiovascular diseases, computed by bivariate LDSC. Connections represent significant *r_g_*, with correlation value along connections, thicker lines denoting stronger correlations, and dark grey denoting more significant correlations. The size of the nodes is weighed by the sample size and *h*^2^_SNP_ of the given phenotype (size = *h*^2^_SNP_ × sqrt(N)).

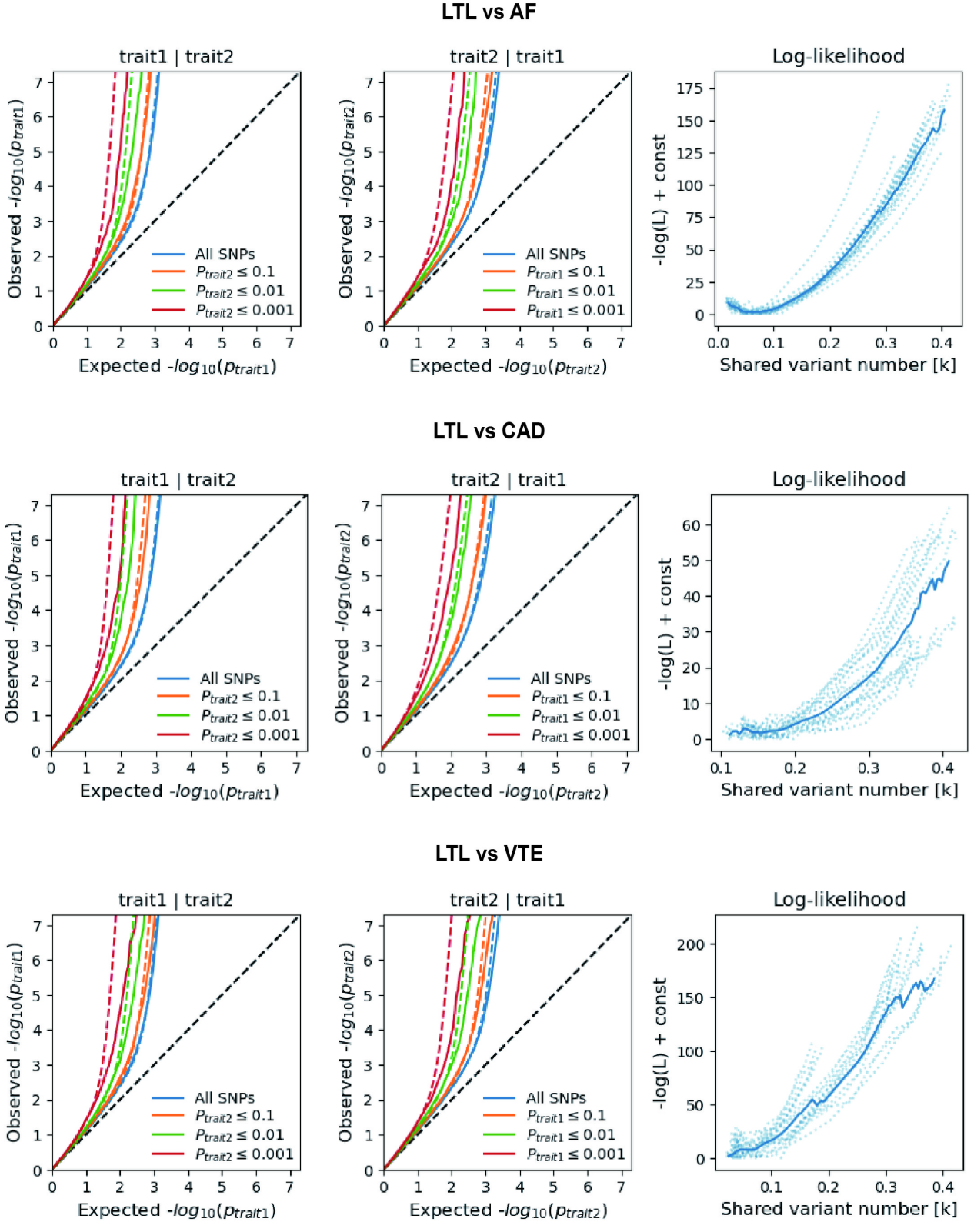

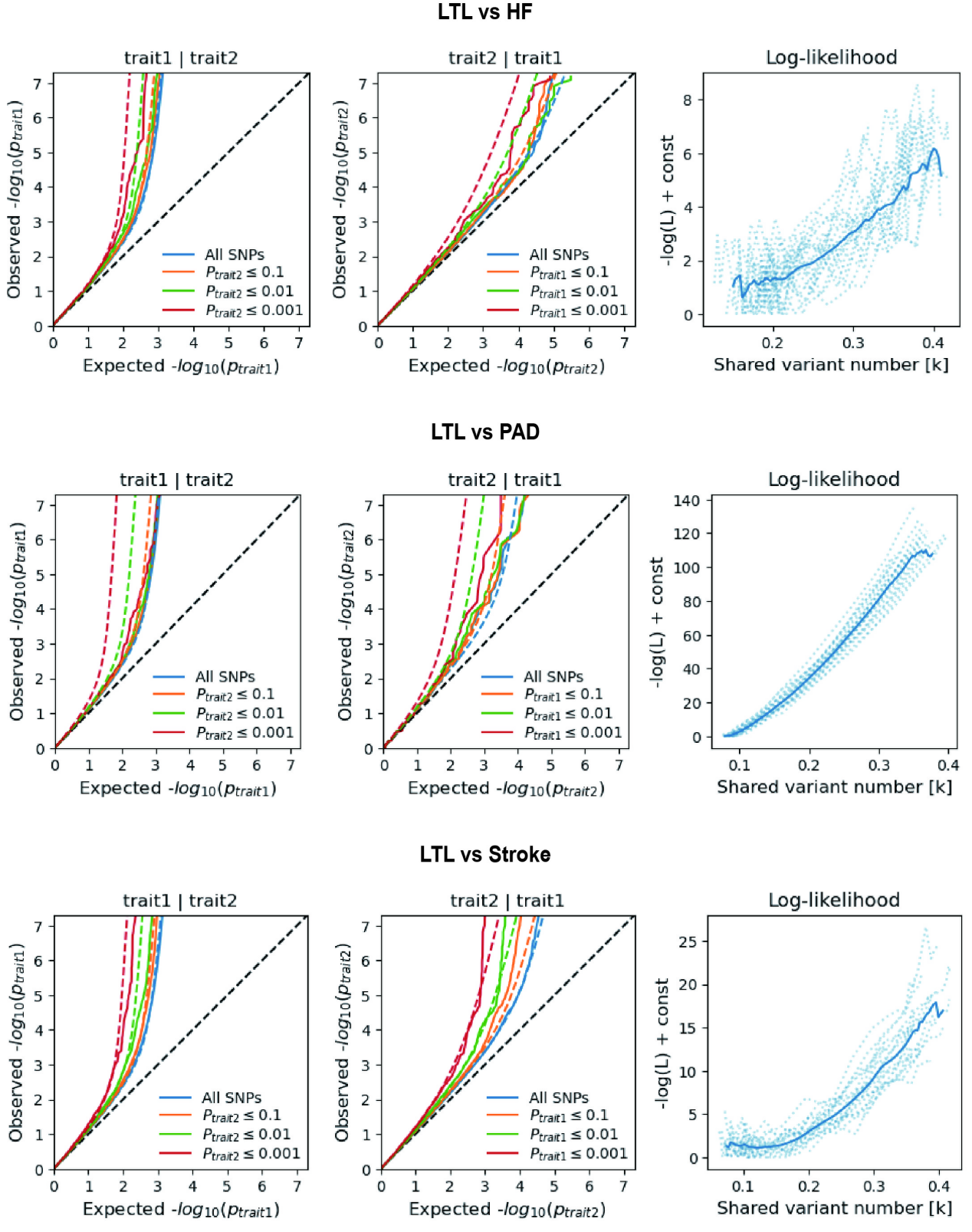

**Supplementary Fig. 2. Supplemental MiXeR figures for each of leukocyte telomere length and six cardiovascular diseases.**

On the left, conditional QQ plots of observed versus expected -log10 p-values in the primary trait as a function of the significance of the association with the secondary trait at the level of all SNPs (blue lines), p ≤ 0.1 (orange lines), p ≤ 0.01 (green lines) and p ≤ 0.001 (red lines). Dotted lines indicate model predictions for each stratum. The black dotted line is the expected Q-Q plot under the null hypothesis (no SNPs associated with the phenotype). Points on the Q-Q plot are weighted according to LD structure, using n=64 iterations of random pruning at an LD threshold r2 = 0.1. On the right, log-likelihood curves highlight the goodness of model fit by plotting the negative log-likelihood function (lower values correspond to better model fit) against the π12 parameter (number of influencing variants shared between two traits). The remaining parameters of the model were constrained to their fitted values. The π12 range on the log-likelihood plots goes from the smallest possible value π12 = *r_g_**sqrt (π1u, π2u) that is still compatible with the estimated genetic correlation, up to the largest possible value π12 = min(π1u, π2u) that corresponds to the minimum total polygenicity among the two traits. The minimum point indicates the best-fitting model estimate of the number of influencing variants shared between two traits.

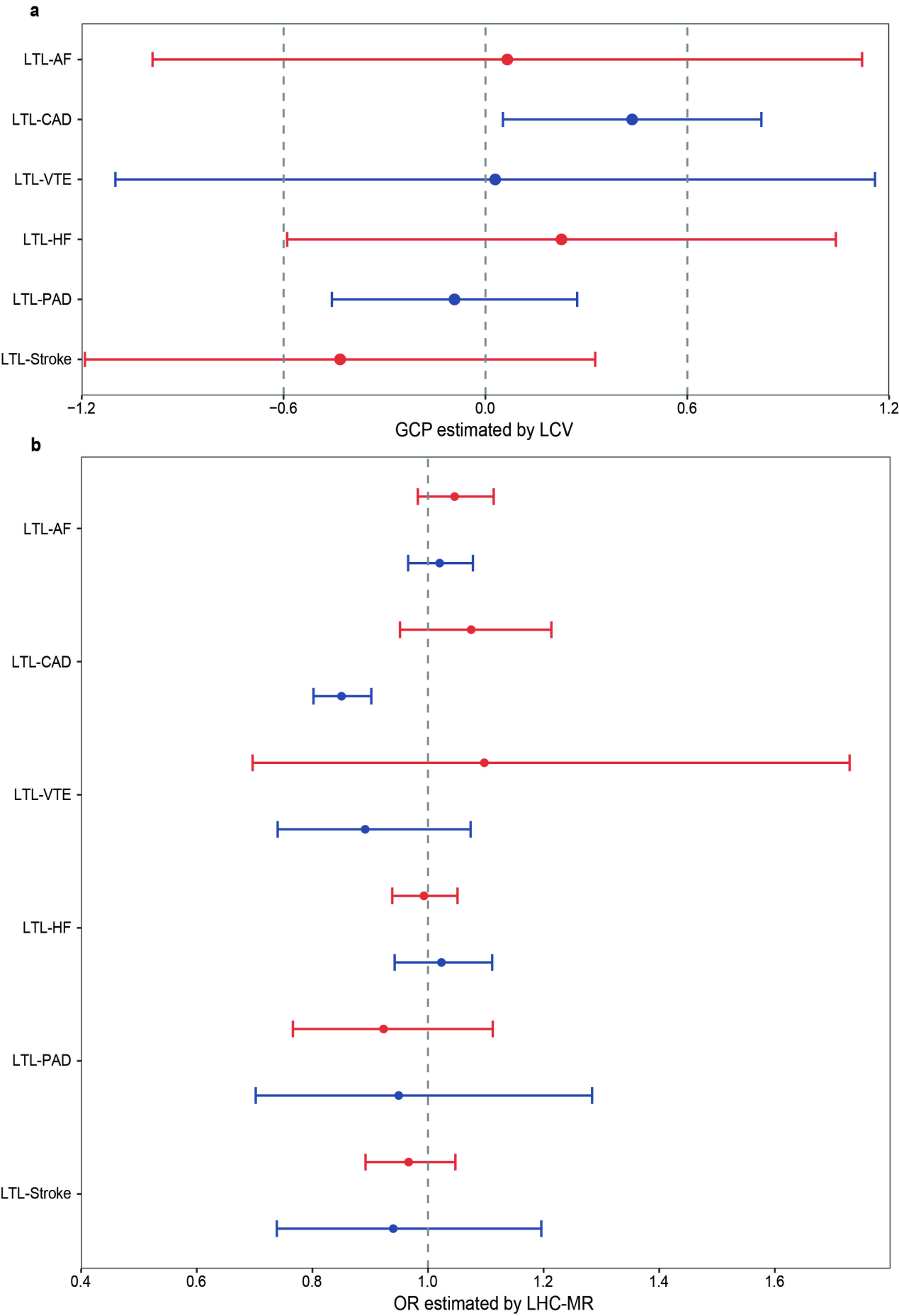
**Supplementary Fig. 3. The causal inference between leukocyte telomere length and six cardiovascular diseases.**

Summary of putative causal relationships between LTL and six CVDs identified by LCV (top) and LHC-MR (bottom). Top: Forest plot of the LCV analysis on the associations between LTL and six CVDs. Circles represent the genetic causality proportion (GCP) estimate, and the error bars indicate standard errors (SE). GCP and SEGCP are posterior mean genetic causality proportions and posterior standard errors estimated by the LCV model. For LCV, a negative GCP indicates a causal effect of LTL on CVDs and vice versa. Bottom: Forest plot of the LHC-MR analysis on the association between LTL and six CVDs. Circles represent the odd ratio (OR) estimate, and the error bars indicate the 95% confidence interval. Results colored in red represent the estimated causal effect of LTL on CVDs, while results colored in blue represent the estimated causal effect of CVDs on LTL. A positive association is indicated by OR > 1, while a negative association is indicated by OR < 1. LTL, leukocyte telomere length; AF, Atrial fibrillation; CAD, Coronary artery disease; VTE, Venous thromboembolism; HF, Heart failure; PAD, Peripheral artery disease.
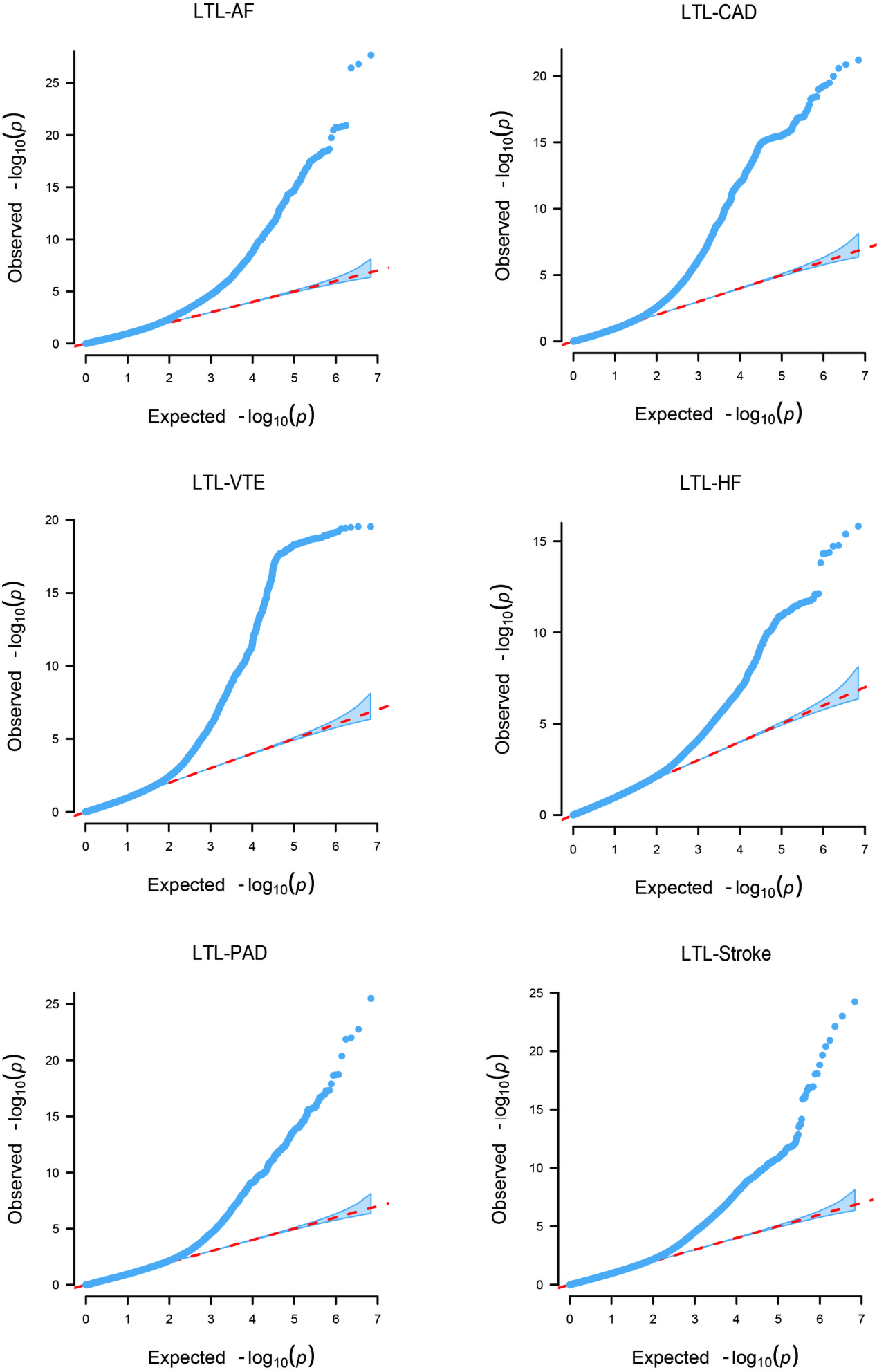

**Supplementary Fig. 4. Quantile-quantile (Q-Q) plots for PLACO results of leukocyte telomere length and six cardiovascular diseases.**

Q-Q plots depicts expected -log10 *P*-values (x-axis) against observed -log10 *P_PLACO_*-values (y-axis). Red dots indicate significant pleiotropic variants (*P_PLACO_* < 5×10^-8^).

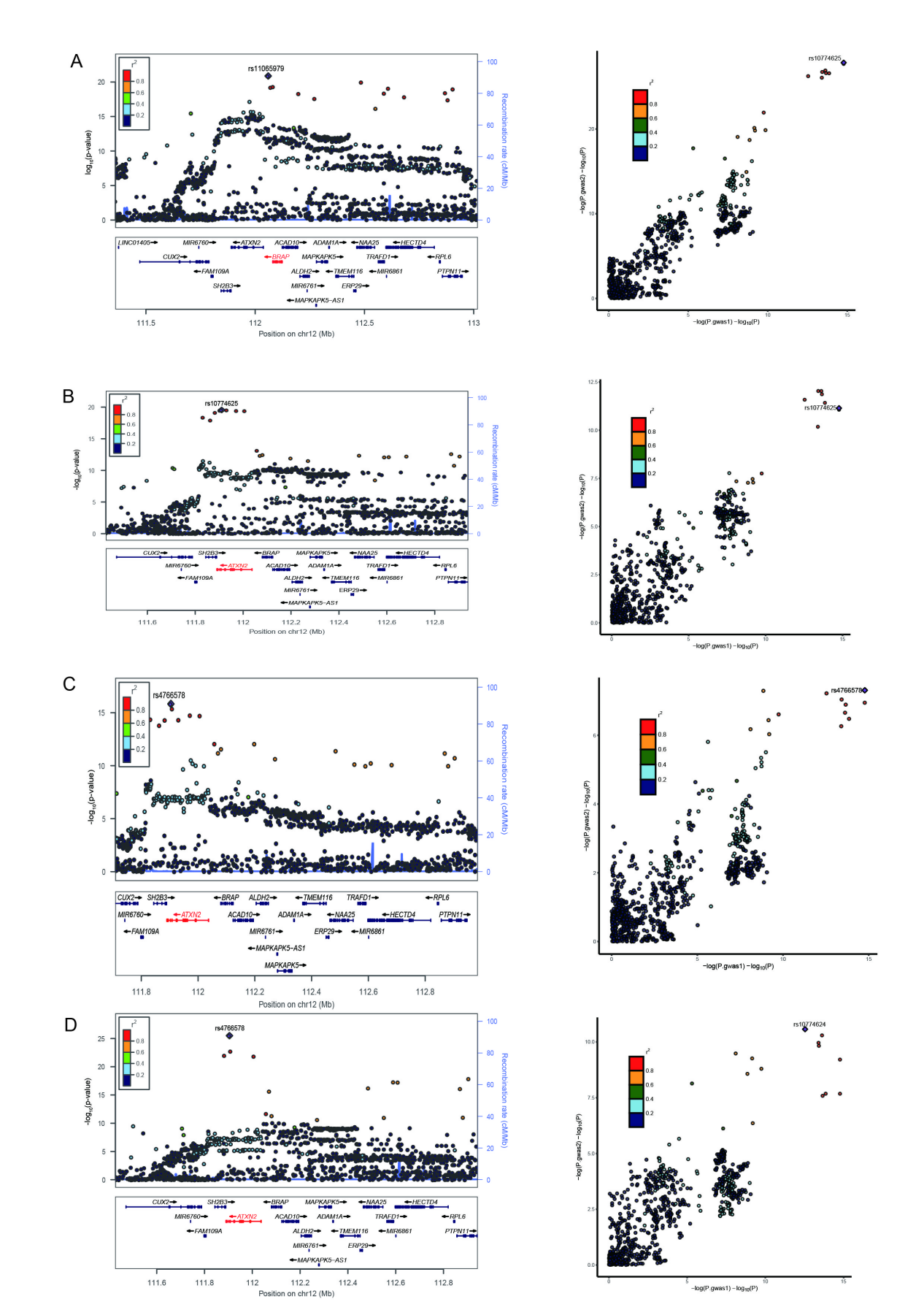

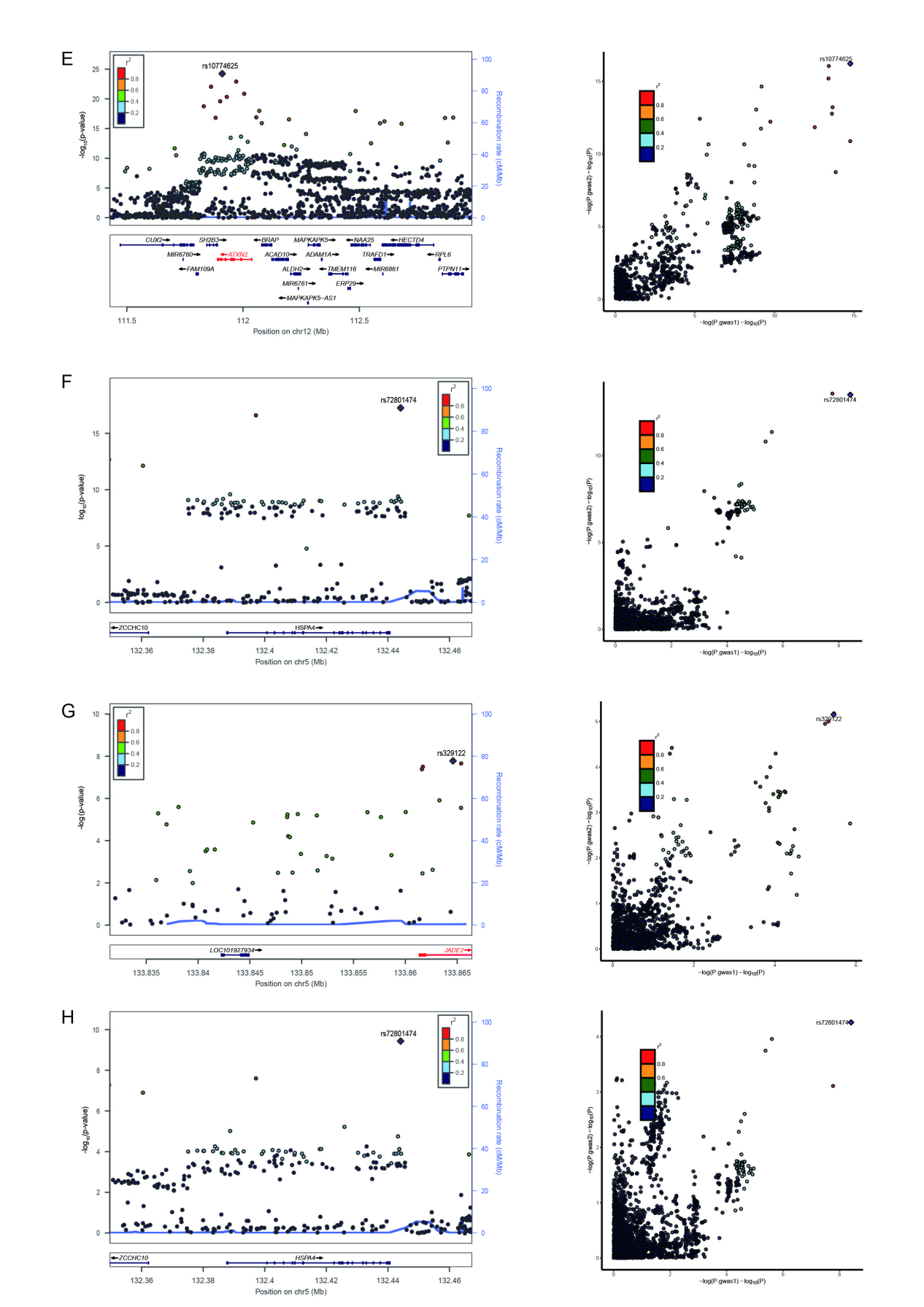

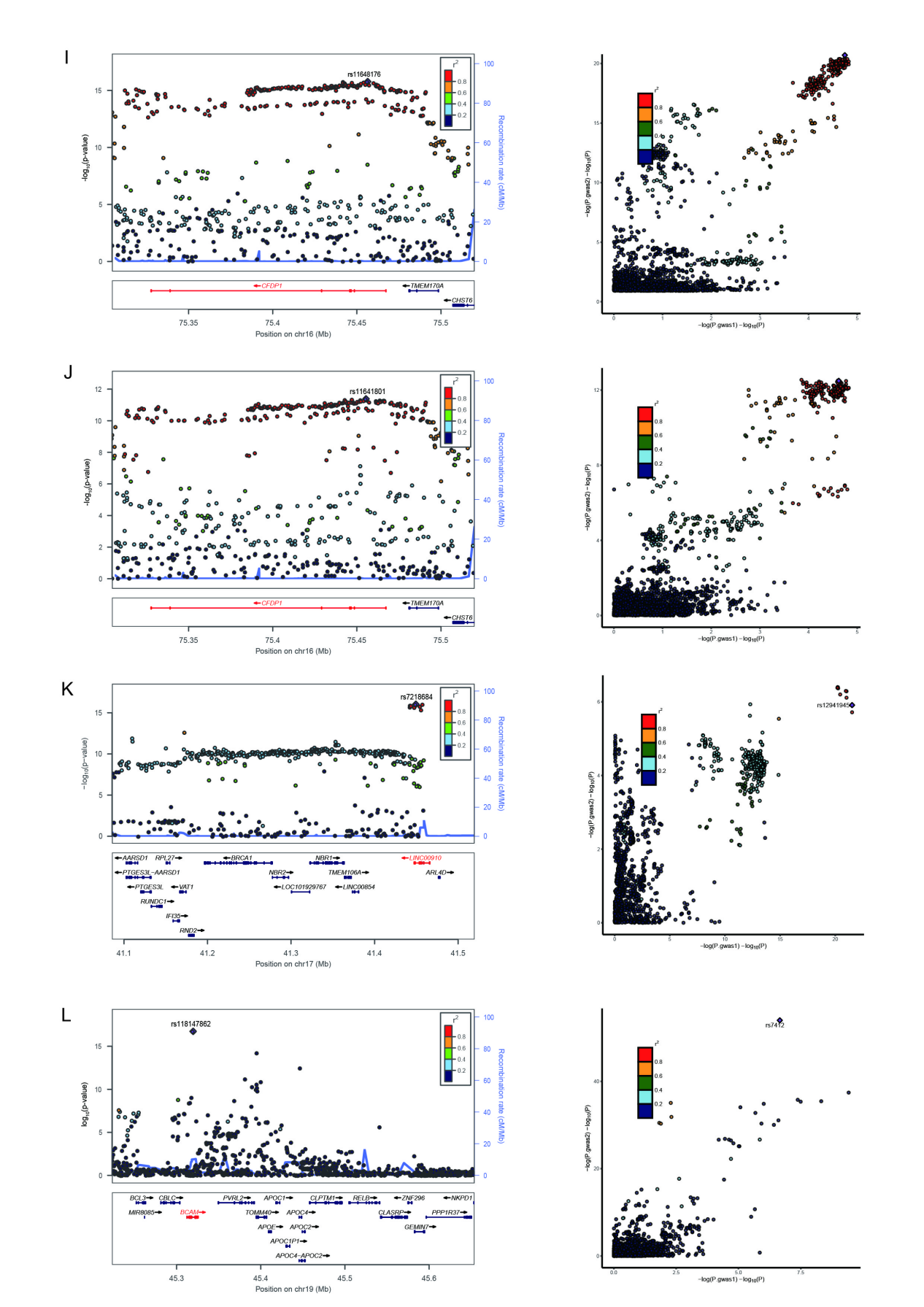

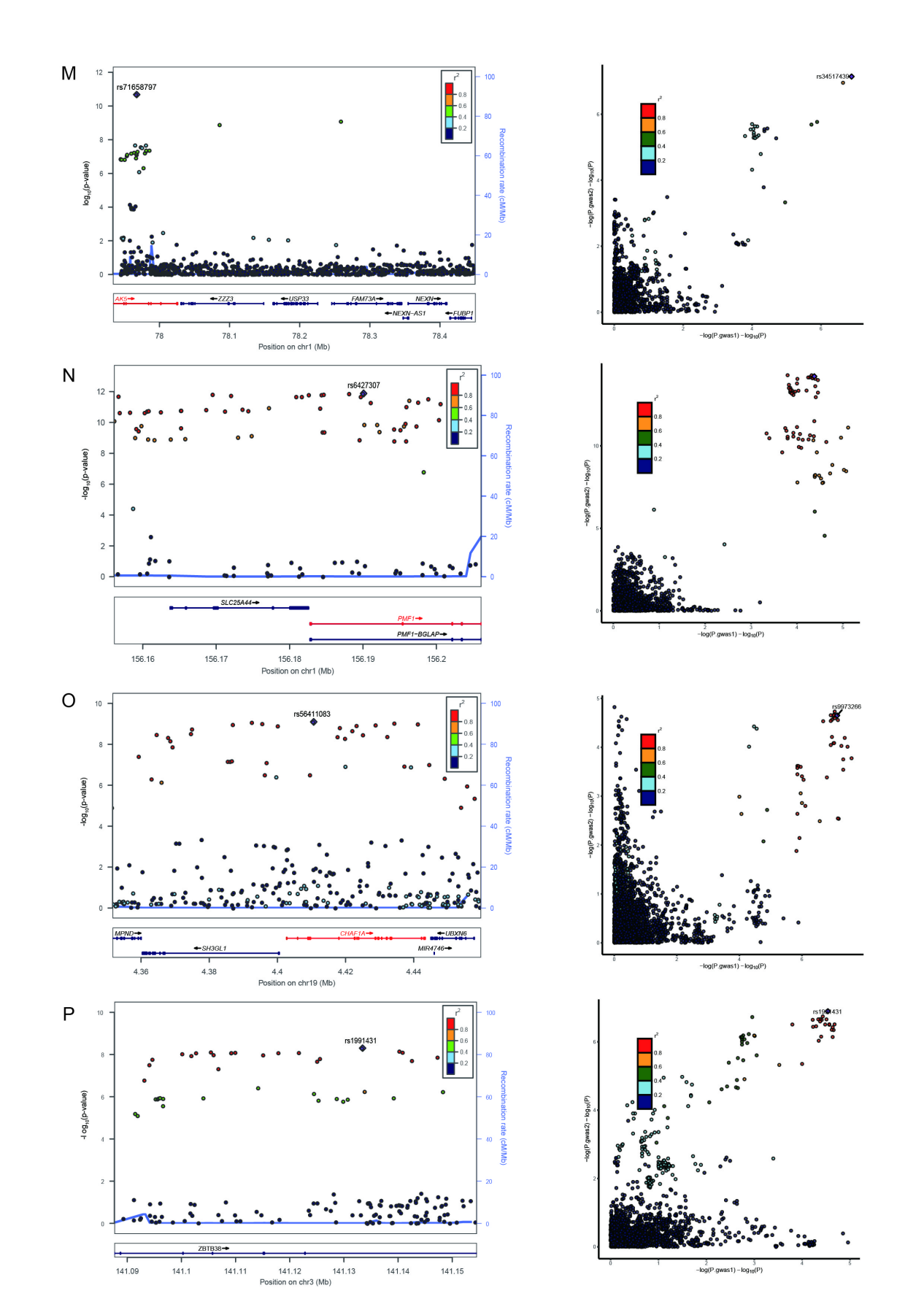

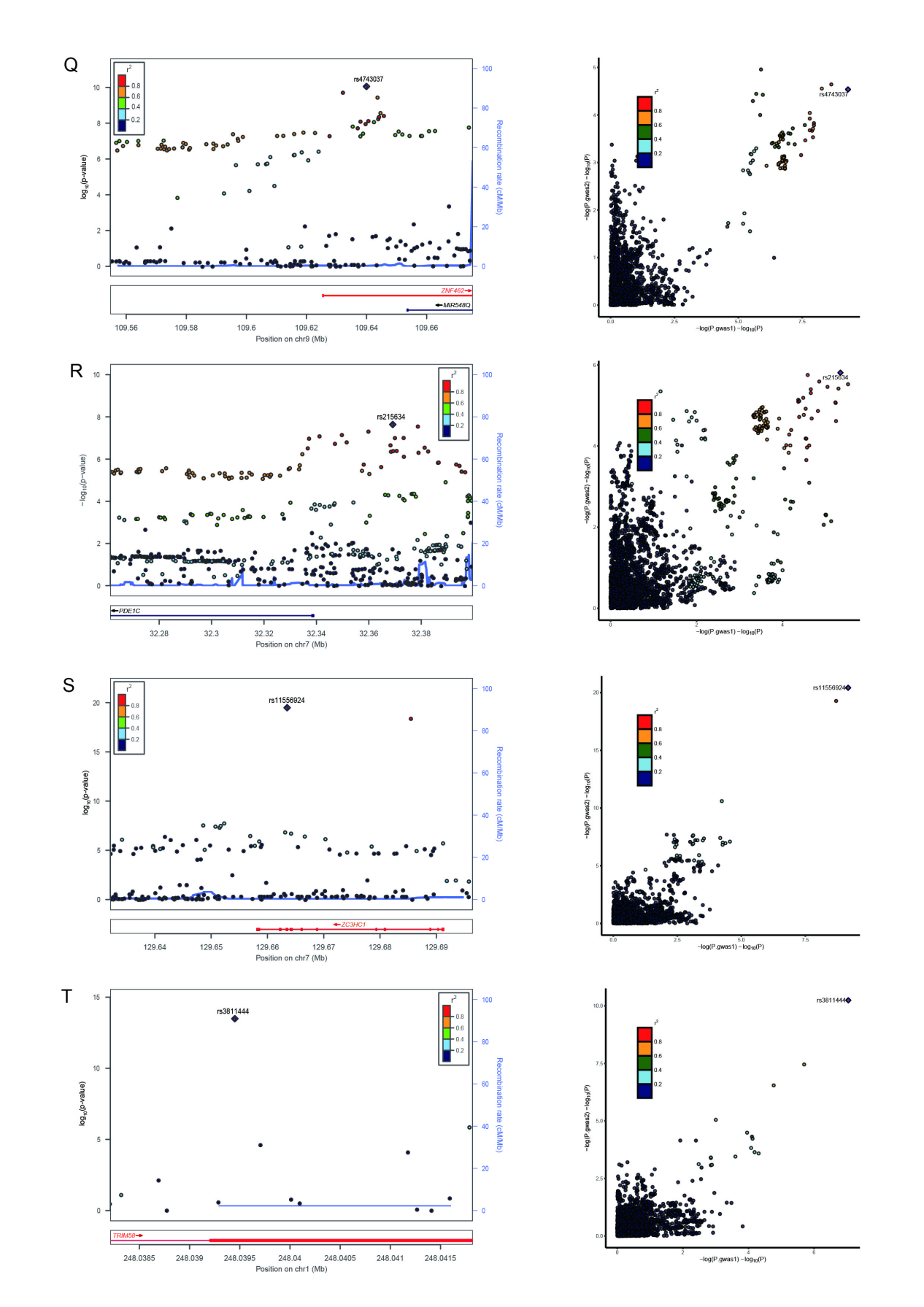

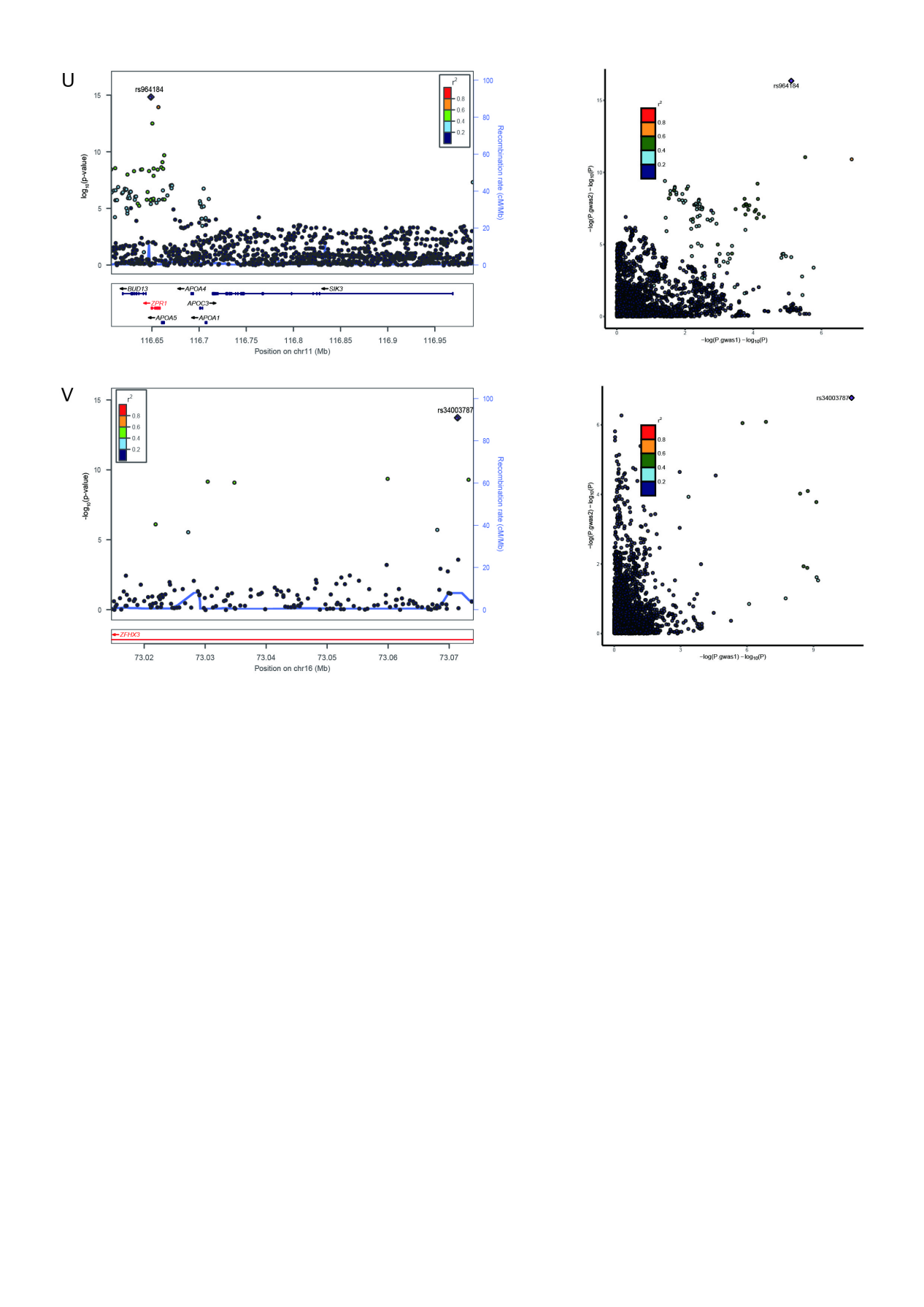

**Supplementary Fig. 5. Locus comparing plots for the shared causal variant for the associations of leukocyte telomere length and six cardiovascular diseases.**

The top 22 genomic loci are influencing LTL and CVDs via shared SNPs. For each colocalized locus (PP.H4 > 0.7) identified for the corresponding trait pair, the left panel depicts the PLACO results using a LocusZoom plot, and the right panel compares two single-trait GWAS statistics of the corresponding trait pair for each variant using LocusCompare plot. For the LocusZoom plot, the x-axis shows the genomic position for each variant, and the y-axis shows -log10 P values from PLCAO results. The top variant with the smallest PPLACO in each locus is indicated in purple diamond. The color of each variant represents its LD relationship with the top variant. For the LocusCompare plot, each dot represents a variant; the x-axis shows the -log10 PGWAS from the corresponding GWAS of LTL, and the y-axis shows -log10 PGWAS from the corresponding GWAS trait. A purple diamond also indicates the candidate-shared causal variant identified by pairwise colocalization analysis. The color of each variant represents its LD relationship with the candidate-shared causal variant. All genomic locations are based on reference genome hg19, and the LD calculation is based on the 1000 Genomes Project of the European population. (A) 12q24.12 for LTL-CAD; (B) 12q24.12 for LTL-VTE; (C) 12q24.12 for LTL-HF; (D) 12q24.12 for LTL-PAD; (E) 12q24.12 for LTL-Stroke; (F) 5q31.1 for LTL-VTE; (G) 5q31.1 for LTL-VTE; (H) 5q31.1 for LTL-Stroke; (I) 16q23.1 for LTL-CAD; (J) 16q23.1 for LTL-VTE; (K) 17q21.31 for LTL-CAD; (L) 19q13.32 for LTL-CAD; (M) 1p31.1 for LTL-VTE; (N) 1q22 for LTL-Stroke; (O) 19p13.3 for LTL-Stroke; (P) 3q23 for LTL-AF; (Q) 9q31.2 for LTL-AF; (R) 7p14.3 for LTL-CAD; (S) 7q32.2 for LTL-CAD; (T) 1q44 for LTL-CAD; (U) 11q23.3 for LTL-CAD; (V) 16q22.3 for LTL-Stroke. Detailed descriptions were provided in Supplementary Table 7.

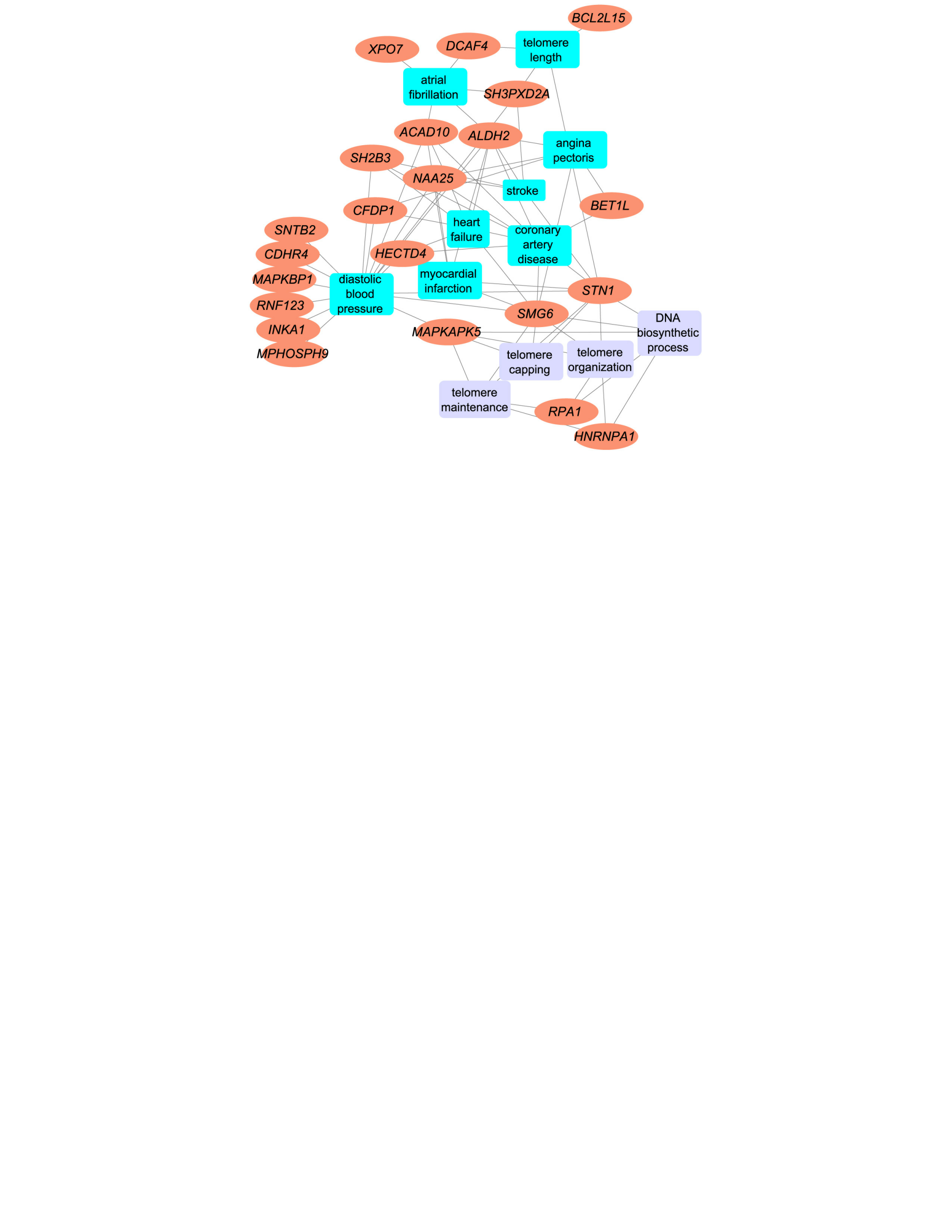
**Supplementary Fig. 6. Supplemental plots for functional enrichment analysis targeting tissue-specific genes using ToppGene Functional Annotation tool.**

A connectivity network that reveals functional features shared to multiple gene lists. The ToppFun functional enrichment analyses (FEA) concerning diseases and pathways validated 22 genes are associated with LTL and CVD-related traits. The four significantly enriched annotations were highlighted, namely ‘telomere maintenance,’ ‘telomere organization,’ ‘telomere capping,’ and ‘DNA biosynthesis process.’

**Supplementary Note 1. Shared genetic architecture contributes to risk of leukocyte telomere length and five major cardiovascular diseases of East Asian.**

**Section 1. Data Sources and Quality Control of East Asian Ancestry**

Analyzing GWAS summary statistics from multiple ancestries reveals a more comprehensive genetic landscape, capturing global diversity and offering an advantage for discovering genetic signals that failed to be identified in European populations. Despite numerous genetic studies on LTL or CVDs in non-European populations, these typically feature smaller sample sizes than those focusing on European (EUR) populations. However, Dorajoo et al. reported a GWAS of LTL in a relatively large Singaporean East-Asian (Southern Han Chinese) ethnic population (n = 23,096)^1^. Both LTL GWAS from UKB and Singapore Chinese Health Study (SCHS) were derived from blood samples collected at baseline and telomere length values were determined using a relative ratio of telomere repeat copy number to a single copy gene (T/S) based on single-color quantitative polymerase chain reaction (qPCR) after adjusting for the technical parameters (i.e., enzyme batch, temperature, operator, primer batch, and humidity). Ishigaki et al. reported a comprehensive GWAS on 42 common diseases conducted through the BioBank Japan Project (BBJ), one of the largest non-European biobanks with approximately 200,000 Japanese individuals. This study included analyses of four major CVDs (CAD, HF, PAD, and stroke)^2^. Concurrently, Low et al. conducted the largest GWAS on AF in the Japanese population, involving 8,180 cases and 28,612 controls^3^. Zhang et al. conducted the first GWAS on VTE in a Han Chinese cohort comprising 1,268 cases and 17,663 controls^4^. As of the latest analysis, GWAS summary statistics for VTE remain unpublished. Finally, we assembled the most extensive available GWAS datasets for five major CVDs in East Asian individuals: AF, CAD, HF, PAD, and stroke. Notably, the sample size for the AF GWAS did not exceed 50,000. Detailed information on these diseases and their publication sources is provided in Supplementary Note Table 1. These GWAS summary data were aligned with the hg19 genome build, referencing the 1000 Genomes Project Phase 3 East Asian. Further details regarding our quality control procedures are elaborated in the main text, ensuring rigorous standards in data analysis.

**Section 2. Genetic overlap beyond genetic correlation between LTL and five major CVDs of East Asian ancestry**

We employed cross-trait linkage disequilibrium (LD) score regression (LDSC) to calculate SNP-based heritability (*h^2^_SNP_*) and to assess genome-wide genetic correlation (*r_g_*) between LTL and five major CVDs. This analysis utilized pre-computed LD scores from the East Asian-based reference panel in the 1000 Genomes Project Phase 3, and SNPs that did not intersect the reference panel were removed from the LDSC analysis. Genetic correlations with *P*-values below the Bonferroni-adjusted threshold (*P* = 0.05 / number of trait pairs = 0.05 / 5 = 0.01) were deemed statistically significant. Univariate LDSC revealed that the estimated *h^2^_SNP_* for LTL was 13.75% (SE = 2.54%). The strength of the genetic signal of GWASs varied considerably among the five CVDs, with the *h^2^_SNP_* ranging from 1.00% to 14.43% (Supplementary Note Table 2a). The estimated *h^2^_SNP_* was higher for AF (*h^2^_SNP_* = 14.43%, SE = 3.15%) and CAD (*h^2^_SNP_* = 6.95%, SE = 0.71%), in comparison to HF (*h^2^_SNP_* = 1.00%, SE = 0.24%), PAD (*h^2^_SNP_* = 1.45%, SE = 0.25%), and Stroke (*h^2^_SNP_* = 1.52%, SE = 0.30%). The results exhibited a notable level of congruity with the findings of the European population. Consistent with findings in European populations, bivariate LDSC analysis detected the most pronounced negative genetic correlation between LTL and PAD (*r_g_* = -0.245, *P* = 0.047, Supplementary Note Table 2b) but fell short of the stringent Bonferroni threshold. No significant genome-wide genetic correlations between LTL and other CVDs were observed, potentially due to the limited sample size and the exclusion of Major Histocompatibility Complex (MHC) regions in the LDSC framework, which may have affected the outcomes.

Although limited genome-wide genetic correlations were observed between LTL and CVDs, this may be attributed to local genetic correlations of opposing directions, which attenuate correlations at the genome-wide level and potentially underestimate the genetic overlap. Additionally, the polygenic nature of LTL could obscure stronger genetic associations with CVD.

MiXeR quantifies polygenic overlap beyond genetic correlation by estimating the total number of both shared and trait-specific causal variants (i.e., variants with nonzero additive genetic effects on a given trait), irrespective of genetic correlation using GWAS summary statistics, and is agnostic to effect directions. Univariate MiXeR revealed that LTL was less polygenic (N = 0.031K, SD = 0.019K). Among the five major CVDs, HF (N = 1.516K ‘causal’ variants explaining 90% of HF’s *h^2^_SNP_*, SD = 0.562K, Supplementary Note Table 3a) demonstrated the highest polygenicity, followed by PAD (N = 0.954K, SD = 0.211K), and Stroke (N = 0.871K, SD = 0.124K). AF (N = 0.029K, SD = 0.007K) and CAD (N = 0.630K, SD = 0.042K) showed lower polygenicity. The large standard deviations for the polygenicity estimates for HF and PAD indicate that these estimates should be interpreted cautiously, likely reflecting a combination of low SNP-heritability and insufficient GWAS power for these disorders.

Bivariate MiXeR results showed that a substantial proportion of the genetic variants associated with LTL also affect CVDs, but the number of shared and trait-specific variants and the balance of

protective and risk-enhancing variants differ across trait pairs (Supplementary Note Table 3b). Causal variants exhibited distinct patterns of polygenic overlap between LTL and CVDs. LTL had similar levels of polygenicity and a more significant degree of polygenic overlap with AF, CAD, and Stroke. For example, we discovered LTL and AF shared 0.005K (SD = 0.002K) out of 0.055K variants with particularly striking polygenic overlap (Dice coefficient  = 0.175, SD = 0.096). The shared variants represent 15.63% of the genetic variants influencing LTL and 16.84% of the variants underlying AF. MiXeR estimated that of the 0.031K LTL-influencing variants, 57.50% and 66.59% also influence CAD and Stroke, respectively. The observed extensive genetic overlap in the Venn diagrams with almost nonexistent genetic correlations suggests a balanced mixture of concordant and discordant genetic effects across shared loci between LTL and these CVD phenotypes (including AF, CAD, and Stroke), with risk variants that did overlap showing low degree of effect direction concordance (34.48%-53.86% of shared variants in the same direction). Given the low polygenicity of LTL compared to highly polygenic diseases such as PAD and HF, substantial differences were observed in the number of shared and unique 'causal' variants. For example, LTL and PAD shared the most significant numbers of ‘causal’ variants (N = 0.031K, SD = 0.019K), with few unique LTL variants (N = 3.22×10^-3^K, SD = 4.29×10^-3^K) and many more unique variants of PAD (N = 0.923K, SD = 0.202K), representing 99.99% of the genetic variants influencing LTL and 3.27% of the variants underlying PAD. While they were moderately correlated at the genome-wide level (*r_g_* = -0.177, SE = 0.036), shared variants were strongly correlated (*r_g_s* = -1.000, SE = 1.16×10^-5^). A similar relationship was evident between LTL and HF. There was complete genetic overlap between LTL and PAD (Dice coefficient  =  0.063, SD = 0.026) or HF (Dice coefficient  = 0.042, SD = 0.024), potentially attributable to the fact that the GWAS of PAD and HF have a substantially more significant effect on sample size than the GWASs of the other CVDs. However, the AIC values were all negative, indicating a poor model fit, and thus, this particular analysis is unreliable.

LAVA estimates local SNP heritability and genetic correlations (loc-*r_g_s*) across 1,445 semi-independent genetic regions of approximately 1.8 Mb. It identifies shared genetic regions with their effect directions despite negligible genome-wide *r_g_*. A total of 124 pair-wise correlations across regions with adequate univariate signals were performed, which yielded 17 genomic regions with nominally significant correlations in at least one trait pair at P < 0.05 (Supplementary Note Table 5). These results provide further evidence of pleiotropy with mixed effect direction among LTL-AF (2 positively correlated and 3 negatively correlated loci), LTL-CAD (3 positively correlated and 2 negatively correlated loci), and LTL-Stroke (2 positively correlated and 2 negatively correlated loci). After correcting for multiple testing using FDR (FDR < 0.05), three out of 17 local genomic regions were found significant for bivariate analysis, and no unique region is shared between LTL with more than one CVD (Supplementary Note Table 5). Only one region identified had a significant negative genetic correlation between LTL and AF (LD block 1,324 on chromosome 18, ranging from 47,731,000 to 51,061,437). Two regions (LD block 912 [chr10:102,471,757-106,139,565] and LD block 1,233 [chr16:25,725,417-27,795,175]) identified between LTL and Stroke exhibited a discordant direction of effect. There were few loci with significant local correlations after FDR < 0.05 due to the smaller sample size and lower heritability of LTL, including no significant loci with CAD, HF, and PAD.

While there was no evidence in this study using the LDSC that significant genome-wide genetic correlations exist between LTL and CVDs, the evidence of extensive genetic overlap verified by MiXeR coupled with almost nonexistent genetic correlation reflects shared genetic etiologies with mixed effect directions, a finding further corroborated by the local genetic correlation using LAVA. Significant local estimates of positive and negative genetic correlation between LTL and CVDs observed for several LD blocks suggest a complex interrelationship between these phenotypes that may exist and warrant additional study.

**Section 3. The causal inference between LTL and five major CVDs of East Asian ancestry**

We evaluated the potential causal relationship between LTL and five major CVDs (i.e., vertical pleiotropy) using LCV analysis. We found strong evidence of a potential causal relationship between LTL and CAD (GCP = 0.676, SE = 0.097, P = 3.83×10^-5^, Supplementary Note Table 6), suggesting that telomere shortening could have a deleterious effect on CAD and implying that their relationship is potentially mediated by vertical pleiotropy. There was insufficient evidence to support a causal relationship between LTL and other CVDs, suggesting the absence of vertical pleiotropy.

**Section 4. Pleiotropic genomic loci identified for LTL and CVDs of East Asian ancestry**

In addition to the usual approaches utilized above, identifying pleiotropic genetic variants or loci responsible for genome-wide genetic correlations enables further exploration of the genetic overlap and shared etiology between these two types of diseases. We employed PLACO to identify genetic variants influencing the risk of LTL and five major CVDs in the same or opposite directions and to explore whether locus-specific effects vary by racial/ethnic group in these regions of genetic overlap. With the smaller sample numbers, East Asian GWAS yielded fewer variants associated with LTL or CVDs than European GWAS. In East Asian individuals, FUMA further delineated 11 independent genomic risk loci as pleiotropic, spanning 7 unique chromosomal regions (Supplementary Note Table 7). Among the loci examined, 8 loci were linked to LTL and 4 loci to CVDs. Notably, a solitary loci showed overlap between LTL and CVDs, accounting for 12.50% and 25.00% of the total loci associated with each category. We found three promising loci shared for multiple trait pairs in 10q24.33, 3q26.2, and 1q42.12. For example, the pleiotropic locus 10q24.33 (mapped gene: *SLK*) was jointly associated with LTL and all CVDs, excluding HF and PAD. The next promising locus is an intron of the leucine-rich repeat containing 34 (*LRRC34*) gene on 3q26.2 encompassing index variant rs9831661 (P = 3.83×10^-11^), and this locus demonstrated associations in LTL-CAD and LTL-Stroke. This variant has been reported to act as a ribonuclease inhibitor in DNA repair, chromosomal stability, and heart development^5^. Most of the top SNPs (81.8%) affected two traits within the trait pair in opposite directions, demonstrating a widespread mixed pattern of allelic effect directions among the shared loci consistent with insignificant genetic correlations. ANNOVAR category annotation across the 11 loci from East Asian ancestry revealed that most index SNPs were intergenic or intronic. Seven of 11 (63.6%) index SNPs were intergenic and three (27.3%) were intronic. No index SNPs with CADD scores greater than 12 were identified, and rs79019201 on 10q24.33 had the highest CADD score (CADD score: 8.428). The four index SNPs had an RDB score of 7, indicating the most minor support for regulatory potential.

Further colocalization analysis showed no polytropic locus had the support of colocalization analysis (PP.H4 > 0.7), reflecting that causal variants for these loci are not effectively tagged in East Asian populations. Two polytropic loci had PP.H3 greater than 0.7, suggesting that different causal variants are implicated in LTL and CVD.

We replicated the shared genetic polytropic loci observed in individuals of European ancestry across trait pairs in those of East Asian ancestry. Further, we discovered loci of genetic overlap exhibiting racial/ethnic differences. Out of the 11 shared loci identified in the East Asian ancestry, 9 of them were also identified in the European ancestry. Only two showed some evidence for being East Asian-specific in that comparison, demonstrating the importance of complementary ancestry-specific loci for identifying associations not shared across diverse populations. Additionally, five loci initially identified in GWAS of AF, CAD, and Stroke in individuals of European ancestry have now been demonstrated to be associated with East Asian populations as well.

**Section 5. Pleiotropic genes associated with LTL and multiple CVDs of East Asian ancestry**

We conducted MAGMA to analyze genes situated within or overlapping the pleiotropic loci, using both PLACO outputs and single-trait GWAS, to identify candidate pleiotropic genes. Our finding of significant gene-level pleiotropy showed 17 potential genes (10 unique), utilizing 22 potential pleiotropic genes located within or overlapping with 11 pleiotropic loci. Among these, 12 genes were detected in two or more trait pairs (Supplementary Note Table 11). *COL17A1* and *SLK* are the top genes identified in 3 trait pairs, followed by *MYNN*, *LRRC34*, and *ACTRT3*, detected in 2 trait pairs. Notably, collagen type XVII alpha one chain (*COL17A1*) and STE20-like kinase (*SLK*) were located on the 10q24.33 locus, identified in all trait pairs except for LTL-HF and LTL-PAD. *SLK* (Ste20-like kinase), a serine/threonine protein kinase, has been identified as a novel kinase that phosphorylates RhoA, a significant regulator of cardiovascular functions. The activation of Rho proteins is a common element in the pathogenesis of hypertension^6^. In addition, through a novel signaling mechanism, *SLK*-mediated activation of the stress response pathway induces senescence by accumulating protein aggregates and causing oxidative and metabolic mitochondrial stress, DNA damage, and pro-inflammatory processes^7^. Modulating the integrated stress response is believed to slow aging and ameliorate age-related pathologies^8^. Of the pleiotropic genes identified, 2 (11.76%) were novel for LTL and 15 (88.24%) for CVDs. No pleiotropic gene was previously reported to be associated with both traits. Furthermore, all genes identified by MAGMA were confirmed by FUMA positional mapping (Supplementary Note Table 9). The number of pleiotropic genes (n = 17) identified in East Asian ancestry was 96.44% less than in European ancestry (n = 478), possibly because the relatively small sample sizes of East Asian samples lead to limited power for predicting genes. Most pleiotropic genes were ancestor-specific, especially, *SH2B3* was specific to Europeans but not included in the East Asians data set, highlighting population-specific disease genetic architecture; three genes (*COL17A1*, *SH3PXD2A*, *STN1*) were generally shared across ancestries.

Overall, both the SNP-level analysis and gene-level analysis in East Asian ancestry converged on the same relevant risk loci, the same potential pleiotropic variants, and the same risk genes.

**Section 6. Shared biological pathways between LTL and five major CVDs of East Asian ancestry**

Shared genetic determinants are likewise mirrored in common biological pathways. Thus, gene set enrichment analysis was also performed to identify potential biological pathways of the significant candidate genes identified from MAGMA. Gene-set analysis revealed that mapped genes for LTL and CAD were enriched in four Gene Ontology (GO) terms, including ‘neutral lipid metabolic process,’ ‘renal system vasculature development,’ ‘glomerular mesangial cell proliferation,’ AND ‘positive regulation of cardiac muscle cell differentiation,’ with a predominance of gene-sets related to positive regulation of cardiomyocyte differentiation (Supplementary Note Table 14). Finally, no significant pathways were identified for AF, HF, PAD, and Stroke. Results from MAGMA’s gene-set analysis using samples of East Asian ancestry were included in comparisons with European ancestry. For most pathways, the enrichment patterns are different, implying potentially different genetic structures for European and East Asian populations.

The absence of a reference panel for East Asian ancestry in specific statistical software, including e-MAGMA, TWAS, and SMR, precluded the identification of tissue-specific genes and causal proteins. Beyond genetic signals shared across populations, our findings suggest the presence of racial/ethnic-specific differences in the pleiotropic effects influencing traits, providing new insights into the shared etiology of LTL and CVD in individuals from East Asian populations. However, we identified only a limited number of lead SNPs, genomic loci, overlapping genes, and gene sets between LTL and five CVDs in East Asian populations, which is not unexpected given the small sample sizes and low power of the original GWAS. This limitation makes it challenging to interpret these shared genetic signals as indicative of a shared etiology of LTL and CVDs. Thus, there is an urgent need for expanded GWAS in the future to deepen our understanding of the genetic architecture underlying LTL and CVDs and putative shared aetiological processes in East Asian populations, which could be significant in targeted therapies and clinical utility.
